# SyMetrics: An Integrated Machine Learning Model for Evaluating the Pathogenicity of Synonymous Variants in the Human Genome

**DOI:** 10.1101/2025.03.21.25324414

**Authors:** Linnaeus Bundalian, Martina Strnadová, Felix Garten, Susanne Horn, Udo Stenzel, Johannes R. Lemke, Saskia Biskup, Björn Schulte, Patrick May, Frank Bösebeck, Antje Garten, Doreen Thor, Angela Schulz, Julia Hentschel, Janet Kelso, Torsten Schöneberg, Diana Le Duc

## Abstract

Synonymous single nucleotide variants (sSNVs), traditionally seen as neutral, are now recognized for their biological impact. To assess their relevance, we developed SyMetrics, a framework that integrates predictors of splicing, RNA stability, evolutionary conservation, codon usage, synonymous variation effects, sequence properties, and allele frequency. We analyzed all possible sSNVs across the human genome, and our machine-learning model achieved 97% accuracy in distinguishing deleterious from benign variants, with a ROC-AUC of 0.89, outperforming individual predictors. Our estimates indicate that about 1.98 ± 0.17% of sSNVs absent from population databases are damaging (roughly 900, 000 sSNVs), with an odds ratio of 3.87 for deleteriousness compared to common sSNVs (p < 0.05). To validate predictions, we performed functional assays on selected sSNVs in the AVPR2 gene. In a clinical cohort, we identified 15 predicted deleterious sSNVs in genes linked to patient phenotypes; 9 were classified as (likely) pathogenic while 6 were variants of uncertain significance (VUS) per American College of Medical Genetics guidelines. For three VUS, segregation data supported their suspected inheritance patterns (de novo, X-linked). Our findings underscore the functional importance of sSNVs. To support further research and clinical applications, we provide a Python package and web application for evaluating these variants comprehensively.

**GRAPHICAL ABSTRACT:** 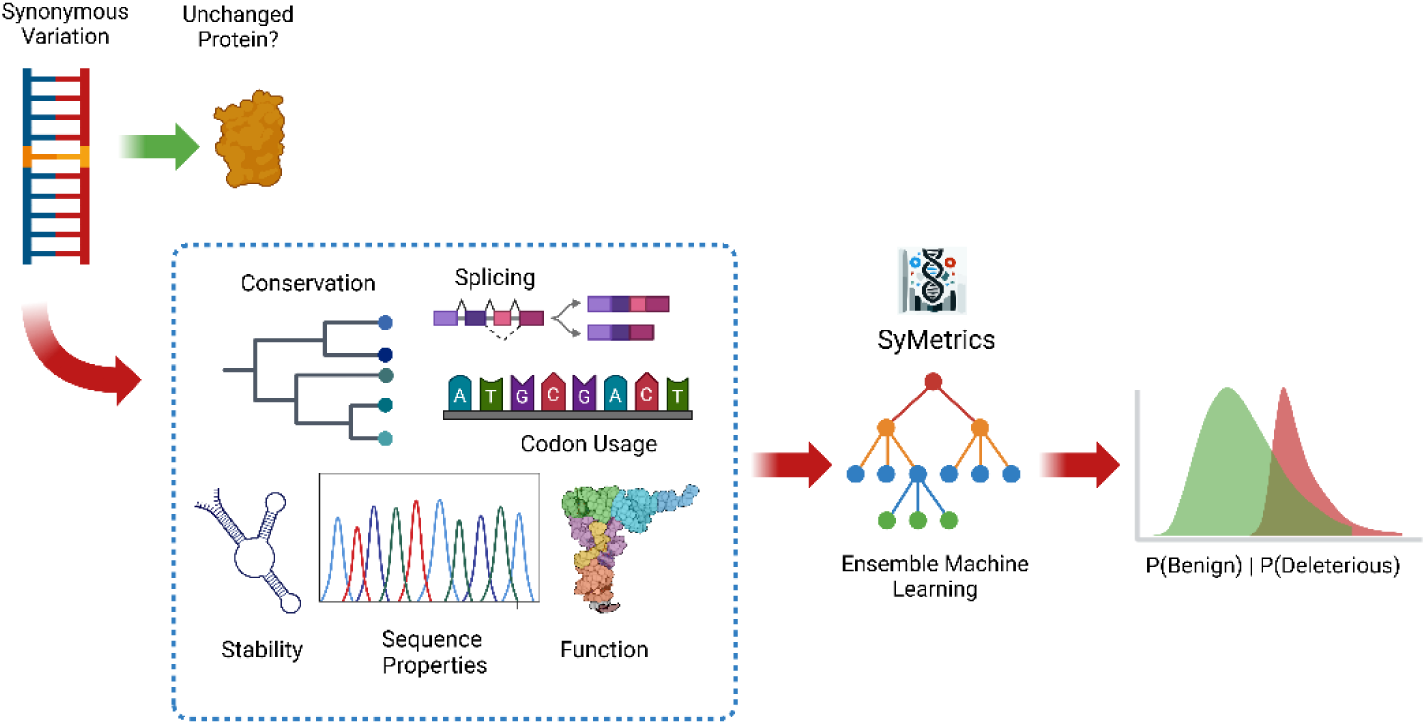

## INTRODUCTION

Recent advancements in genomics and molecular diagnostics have significantly enhanced our understanding of genetic variation. Emerging sequencing technologies have provided invaluable resources for research, enabling deeper exploration of the human genome (1, 2) and greater insights into the mechanisms underlying various diseases and complex traits (3). However, despite substantial progress in diagnostic yield (4), a significant diagnostic gap remains, particularly in rare diseases, with approximately 50% of cases still undiagnosed (5, 6).

Single nucleotide variations (SNVs) constitute a major component of human genomic variation and have drawn considerable attention from the medical and research communities. An SNV involves a single nucleotide change (7), which can be classified as either synonymous (sSNV) or non-synonymous (nSNV). Synonymous SNVs (sSNVs) do not alter the amino acid sequence during translation (1, 3, 8–13) and are often referred to as silent mutations (1, 10). In contrast, non-synonymous SNVs (nSNVs) change the protein sequence and potentially its function, making them the primary focus of disease and trait-related studies (8, 14).

Traditionally, human genetics, mutagenesis screens, and evolutionary analyses have suggested that sSNVs are more likely to be neutral compared to nSNVs (15). However, a growing body of research challenges this assumption, with studies indicating that sSNVs can have significant functional consequences (15, 16). Recent findings suggest that purifying selection, a form of natural selection that removes harmful genetic variants, also acts on synonymous mutations (17), reinforcing the idea that they are not necessarily neutral. Even without altering amino acid sequences, sSNVs can impact gene expression and protein function through mechanisms such as modifying mRNA stability, altering splicing efficiency, and affecting translation kinetics (13).

Furthermore, the human genome contains alternative open reading frames (ORFs) embedded within canonical mRNAs (18, 19). Although these alternative ORFs are less well-annotated and often exhibit lower expression levels, they add an additional layer of complexity to genetic interpretation. Notably, an sSNV in a canonical transcript may result in a non-synonymous change when translated in an alternative reading frame, further highlighting the functional potential of sSNVs.

Given this complexity, annotating phenotype-affecting variants as “synonymous” may be misleading in clinical settings. To address this, it has been proposed that functionally consequential sSNVs be termed “unsense” mutations for more precise communication (20). However, existing methods for quantifying variant effects are largely tailored to nSNVs (21, 22), with few tools specifically designed for assessing sSNVs. In response, several synonymous variant effect predictors have been developed, including Silent Variant Analyzer (SilVA) (23), Identification of Deleterious Synonymous Variants (IDSV) (10), and others (24). These initial efforts have demonstrated the need for robust predictors and have driven further interest in developing and benchmarking bioinformatic tools for synonymous variant interpretation (24, 25).

In this study, we sought to understand how sSNVs disrupt biological processes or impair gene function. To develop a predictive framework, we integrated existing functional consequence scores (synVEP), splicing effect predictors (MES, SpliceAI), RNA stability and folding metrics (SURF), phylogenetic conservation scores (GERP++), codon usage measures (RSCU, dRSCU), and sequence properties (e.g., CpG sites, CpG exons, and proximity to mRNA and pre-mRNA). To classify sSNVs, we trained an ensemble machine learning model using a dataset of known benign and deleterious synonymous variants (26), incorporating synonymous variant effect measures and allele frequency as features. Our model estimates that approximately 1.98 ± 0.17% (95% CI) of synonymous variants absent from population databases are deleterious. Functional validation demonstrated that predicted deleterious variants in *AVPR2* impair function. Finally, we reanalyzed a large clinical cohort, identifying potentially deleterious sSNVs in genes associated with the probands’ phenotypes. To facilitate research and clinical applications, we provide the results through the Python library and web platform – SyMetrics (tools.hornlab.org/symetrics), enabling researchers and clinicians to differentiate deleterious sSNVs from benign ones.

## MATERIALS AND METHODS

### ‘OBSERVED and ‘NOT SEEN’ Synonymous Variants

The sSNVs in the synVep or generated database were classified as follows:

- ***observed* –** variants which can be found in the gnomAD database with allele count greater than 1 (AC > 1)
- ***not seen*** – variants which cannot be found in gnomAD database.
- ***singleton*** - variants which can be found in the gnomAD database but with AC = 1.
- ***unobservable*** - variants which are not likely to be observed in the future were discarded in the previous study as it deemed to have little to no effect in the analysis.

We regrouped the variants by putting both ***singleton*** and ***observed*** together as **observed** and ***unobservable*** and ***not seen*** as **not seen** while using only the variants which passed the QC filter of latest gnomAD version (v4).

### sSNV Effect predictor, Features and Scores

There are several existing tools and effect predictors designed to target sSNV (21, 22, 27). We used SynVep, SpliceAI, SURF RNA stability, and SILVA to generate scores. For SILVA, we focused on the intermediate output of the model, as these are easier to explain and relate to biological implications and relevance. These tools helped us acquire scores reflecting the sSNV’s metrics related to its consequences, splicing effects, RNA stability, phylogenetic and conservation scores, codon usage, and sequence properties, as described in Supplementary Table 1 – Model metrics.

#### Functional Effect – synVEP

One of the recently developed tools for assessing the functional impact of synonymous variants is the machine learning-based tool synVep. This tool assigns scores to sSNVs - synVEP scores - by using a sequential extreme gradient boosting model to differentiate pathogenic from benign variants. The score generated reflects the potential impact of the sSNVs. The implications of the scores are shown in the following notation:

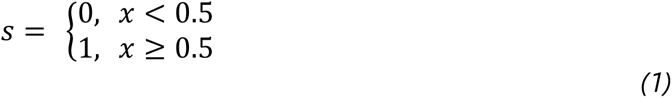

Where 0 = no effect and 1 = effect (25).

#### Splicing Effect – SpliceAI and MES

One of the most important aspects that essentially affects gene activities as well as protein diversity is splicing – the process involving the removal of introns to join exons together hence forming the mature messenger RNA and disrupting this process can lead to aberrant production of mRNA which can manifest as disease or unexpected phenotypes (28, 29). SpliceAI is a deep learning tool developed to predict the probability of whether a given position of pre-mRNA sequence is a splice acceptor or donor site in terms of Delta Scores (Δ Score) for acceptor loss, donor loss, acceptor gain and donor gain (30). High scores, at least greater than 0.22 (31), are indicative that the position has a higher probability of being a splice acceptor or donor site. However, threshold may vary depending on the context. In this study, we used the maximum delta scores (Max Δ Score or MAX_DS) among the four delta scores for a given genetic variant to simply indicate that the variant indeed can cause splicing. Additionally, we used MES generated from MaxEntScan to estimate the splice site motif or junction strength (32, 33). The values represent the maximum efficiency of splicing at the sites. We used MES ≥ 3 as a threshold to indicate a relevant and potential influence of the genetic variation to splicing. The established threshold was based on the empirical observations on overall and combined score distribution of both observed and not-seen and is best to represent at least the top 2% of the population.

#### RNA Stability – SURF

Summarized RNA Folding (SURF) is another metric under consideration that focuses on how genetic variants, especially sSNVs, affect RNA stability and folding. It calculates a broad range of RNA folding metrics for each SNV and evaluates the changes induced by this genetic variation, including, but not limited to, free energy, edge distance, and centroid distance. From these metrics, a unified score is derived to represent the impact of the variants under observation. For our analysis, we used a SURF threshold of ≥ 5 to indicate that the variant may have a potential influence on RNA stability (34).

#### Conservation and Phylogenetic Relevance - GERP++

The evolutionary constrained elements were estimated using GERP++ which is a valuable tool capable of quantifying the level of evolutionary constraint which can be indicative of their functional influence and selection pressure. This tool calculates site-specific rejected substitution scores (RS) where higher positive scores imply more deleterious genetic variants and/or stronger selection(35) similar to its predecessor GERP (36). The threshold is set as GERP++ ≥ 4, which is considered to have a large deleterious effects associated with a high selection coefficient (37–39).

#### Codon Usage

Two essential metrics used for codon usage analysis are RSCU (Relative Synonymous Codon Usage) and dRSCU, which represent the standardized differences in RSCU values. RSCU assumes that all synonymous codons are used equally, and based on this assumption, it compares the observed and expected frequencies of codons. A value greater than 1 implies overrepresentation, while a value less than 1 indicates underrepresentation (40, 41). Therefore, for this score, we established two criteria to observe the outcomes: one for when the threshold is RSCU > 1 and another for RSCU < 1. On the other hand, dRSCU helps assess the deviation of specific codons from common usage patterns in a dataset (42). The range of observed values for dRSCU is from 0 to 2, and we decided to set the threshold at dRSCU > 1, as we aim to focus on values that are significantly different from 0, where there are minimal or no observed standard differences.

#### Sequence Features

Although nucleotide sequences may appear complex and unintuitive at first glance, closer examination reveals recurring patterns that are associated with molecular signatures and biological processes (43, 44). We used CpG and relative distance to pre mRNA (f_premrna) and mature mRNA (f_mrna). CpG sites are DNA regions where cytosine (C) and guanine (G) and the overall occurrence of CpG contents provides a baseline reference with which actual observations can be compared. Significant deviations from the expected CpG might suggest functional relevance. The features f_mrna and f_premrna capture the relative position of the genetic variant in a mature and pre mRNA transcript (45).

### Gene-wise Scoring Function and Constraints Group Comparison

With the listed effect measures, we aimed to identify signals that could be induced by different features related to these measures. This approach provides insight into the relevance of the features as well as the effectiveness of the selected scoring scheme. Specifically, we determined which genes had more sSNVs with high scores, i.e., where the score for a particular effect measure exceeded the set threshold, in relation to the proportions of the observed and not seen groups. To assess the difference in proportions between these groups for each given effect measure/scoring scheme, we used a z proportion test.

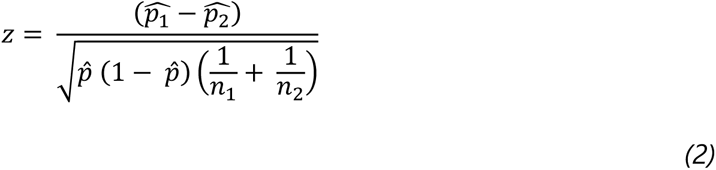

Where:

- *p̂* – the pooled proportion of variants meeting the given criterion and score threshold for both NOT SEEN and OBSERVED
- *p̂*_1_ – the proportion of NOT SEEN variants meeting a given criterion and score threshold.
- *p̂*_2_ – the proportion of OBSERVED variants meeting a given criterion and score threshold.
- *n_1_* – the number of NOT SEEN variants.
- *n_2_* – the number of OBSERVED variants
- *z* – the resulting statistic from the proportion test as a function of the selected/given variant effect measure (*synonymous Z-score*).

Using the proportion also allows us to account the length of the genes since with this approach we normalized the aggregated value by the number of observed values. A synonymous Z-score means that there are more not seen variants having a high effect measure score compared to the observed variants and lower score means otherwise.

The variant effect and gene-wise scores can aid in the deterministic interpretation of the results. The combined implications of the scores can be represented as follows:

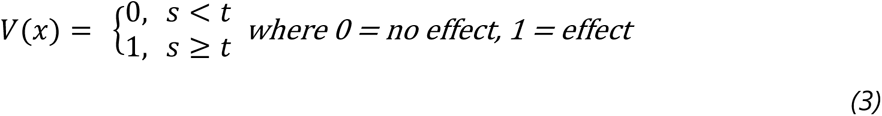

Where:

- *t* – is the set threshold for the selected effect measure
- *s* – is the effect measure score
- *V(x)* – is whether that score implies that the variant has an effect or none.

And,

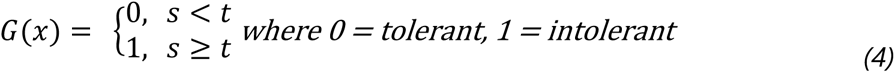

Where:

- *t* – is the set threshold for the selected effect measure
- *s* – is the gene-wise score
- *G(x)* – whether the score implies that the gene is tolerant or not.

### Functional Implications and Enrichment Analysis

To further interpret the scores, we attempted to relate them to existing gene constraints in gnomAD - specifically, *pLI* and *Missense Z scores*. After scoring the genes, we grouped them into categories based on whether they had HIGH or LOW scores in the selected gene constraints. This categorization was done to determine if genes with high gene constraints tend to have higher gene-wise scores from the previous step. The group definition is as follow:

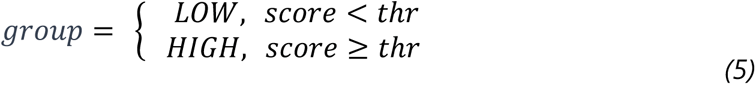

Where:

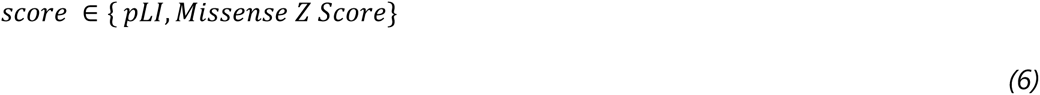

To test in which functional pathway the genes identified to have a high gene-wise score play a role, we performed enrichment analysis and overrepresentation analysis across different gene sets in Gene Ontology (GO), KEGG and Reactome using the R package clusterProfiler (46, 47) which uses hypergeometric test (48) to assess the probability of observing at least *k* genes from the list of the pathway database.

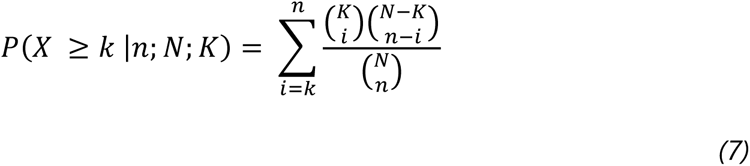

### Variant Classification Model

#### Dataset for known deleterious and benign sSNV

The known disease-causing dataset, representing the positive class, was retrieved and benchmarked from an existing study on synonymous variants (26). The list consisted of variants from the Database of Deleterious Synonymous Mutations (dbDSM v1.2) (49) and the ClinVar database, with variants labeled as ‘likely pathogenic’ and ‘pathogenic’ considered as true positives. The negative class was represented by variants labeled as ‘likely benign’ and ‘benign’ in ClinVar (26, 50). The pooled dataset included 4, 696 benign sSNVs and 367 deleterious sSNVs, all mapped to the GRCh37 reference genome. For model training and validation, the dataset was split into 70% for training, 15% for testing, and 15% for validation, while Synthetic Minority Oversampling (SMOTE) was used to address the class imbalance. This method augments the minority class by randomly selecting an instance and finding its k-nearest neighbors, generating a new instance that is a convex combination of the selected instance and its nearest neighbors. This helps the model build larger decision regions around the minority class points without compromising data integrity (51). We verified the integrity of the resampled data by checking the distance between clusters formed from the resampled data and their original class, confirming that the resampled clusters were near the original ones (Supplementary Fig. 4).

### Model Selection and Evaluation

For the ease of model selection, we used the Python library – Lazy Predict (52). This library automates the training and evaluation of multiple classification models from the most traditional up to the advanced methods. The output does not give the models themselves but rather yields the metrics of the baseline models that can aid in the selection. Upon selection, it is still necessary to train it in the main library supporting the selected model from the Lazy Predict module. The following metrics and/or statistical measures (Table 1) were used to evaluate the performance of the models.

**Table 1.**
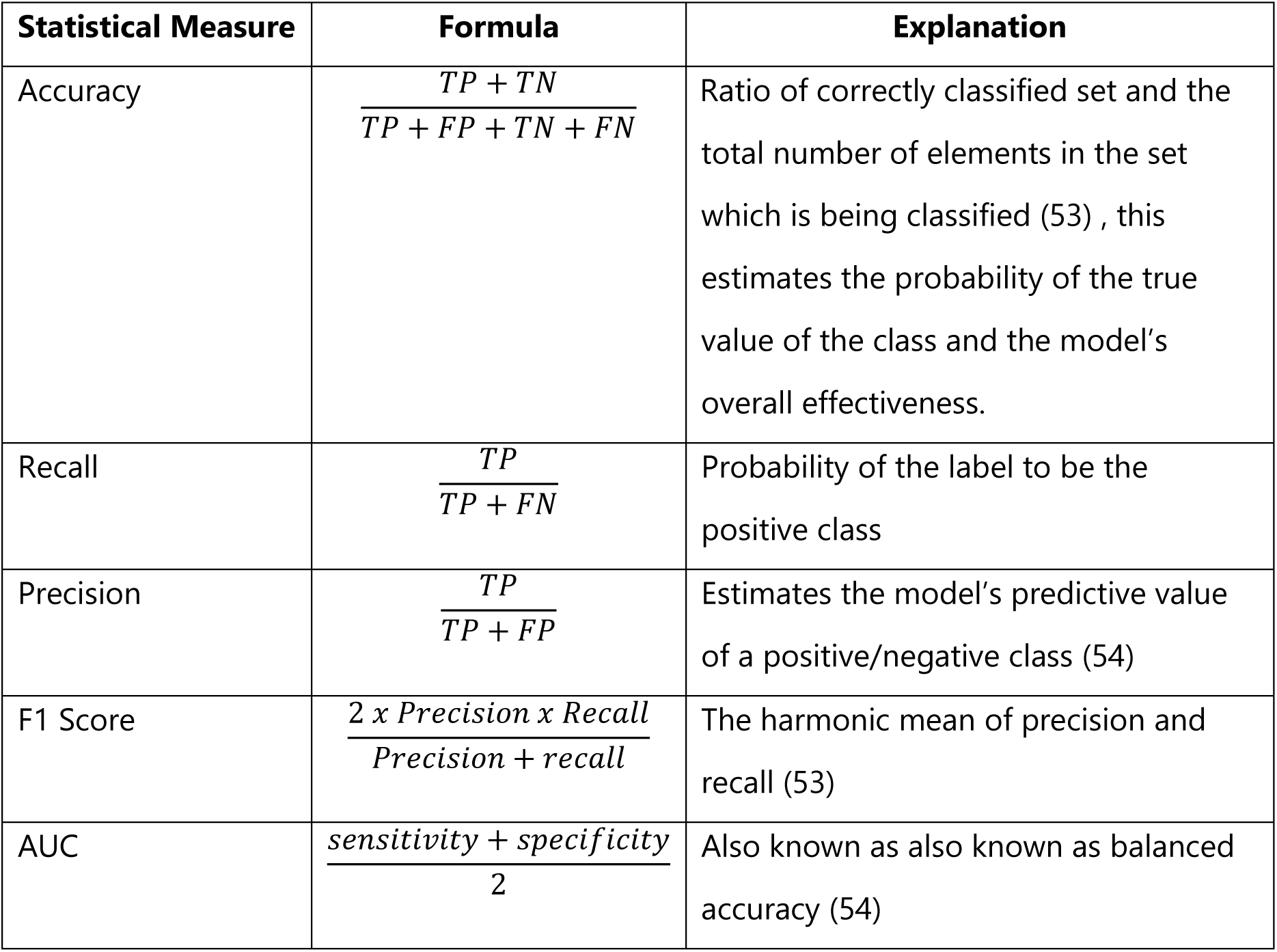
Performance metrics are used to evaluate the models.

| Statistical Measure | Formula | Explanation |
| --- | --- | --- |
| Accuracy | $\frac{TP + TN}{TP + FP + TN + FN}$ | Ratio of correctly classified set and the total number of elements in the set which is being classified (53) , this estimates the probability of the true value of the class and the model's overall effectiveness. |
| Recall | $\frac{TP}{TP + FN}$ | Probability of the label to be the positive class |
| Precision | $\frac{TP}{TP + FP}$ | Estimates the model's predictive value of a positive/negative class (54) |
| F1 Score | $\frac{2 \times Precision \times Recall}{Precision + recall}$ | The harmonic mean of precision and recall (53) |
| AUC | $\frac{sensitivity + specificity}{2}$ | Also known as also known as balanced accuracy (54) |

**Table 2.** Model Comparison of Lazy Predict Top 5 Models. The selection of the best model is based on their performance on accuracy, precision, recall, F1 Score, and ROC-AUC. Note that emphasis on the balance of precision and recall should be considered along with accuracy. (Note: ExtraTrees – Extremely Randomized Trees, LGBM – Light Gradient Boosting Machine, Nu SVC – Nu Support Vector Classification, XGB – Extreme Gradient Boosting)

| Model | Accuracy | Precision | Recall | F1 Score | ROC AUC |
| --- | --- | --- | --- | --- | --- |
| **Extra Trees** | 0.963 | 0.814 | 0.636 | 0.714 | 0.879 |
| **LGBM** | 0.958 | 0.750 | 0.627 | 0.683 | 0.873 |
| **Nu SVC** | 0.882 | 0.338 | 0.664 | 0.448 | 0.831 |
| **XGB** | 0.955 | 0.725 | 0.600 | 0.657 | 0.881 |
| **Random Forest** | 0.953 | 0.683 | 0.645 | 0.664 | 0.878 |

### Symetrics Web and Stand-alone Package

We created a python package (https://pypi.org/project/symetrics) and a web platform (https://tools.hornlab.org/symetrics/) to allow developers, clinicians, and researchers to access the records of the synonymous variants along with the selected variant effect measures as well as the predictions.

### Functional Assay and Analysis of Selected Synonymous Human Genetic Variant Materials

If not stated otherwise, materials used for cell culture were obtained from Sarstedt or ThermoFisher. Standard chemicals were purchased from Merck or Carl Roth GmbH. Primers were synthesized by Microsynth.

#### Cloning and introduction of point mutation

The genomic sequence of human *AVPR2* was amplified using human genomic DNA. Thereby, an N-terminal hemagglutinin (HA)-Tag and a C-terminal FLAG-Tag were introduced after start codon and in front of the stop codon, respectively. Both tags have been shown to not interfere with receptor expression and function (55, 56). Furthermore, the amplification primers:

- forward primer: 5’- TGTACCCCTACGACGTCCCCGACTACGCCCTCATGGCGTCCACCACTTCCG-3’
- reverse primer: 5’- ATCATGTCTGGATCCACTAGTCACTTATCGTCATCGTCCTTATAATCCGATGAAGTGTCCTT GGCC-3’

contained restriction sites for *Aat*II and *Spe*I to allow for cloning into the pcDps-derived plasmid pL (57). To insert the point mutations, quick change mutagenesis was performed. Thus, two complement primers were designed carrying the desired point mutation in the middle of the primer sequence (Supplementary Table 1 − Primers used to insert point mutations). PCR was performed using Phusion high fidelity polymerase (ThermoFisher) according to the manufacturer’s protocol using AVPR2 pL as template plasmid at an annealing temperature of 55°C. Prior to transformation into chemical competent cells, template plasmid was digested using *Dpn*I (NEB). Sanger sequencing was performed by Microsynth to ensure the correct sequences of the mutants derived.

#### Cell Culture

HEK273T cells were cultured in DMEM media (ThermoFisher) supplemented with 10% fetal bovine serum (FBS), 100 U/ml penicillin and 100 µg/ml streptomycin at 37°C in a humidified atmosphere containing 5% CO_2_. Cells were regularly split twice a week. For the different analysis, cells were counted using a Neubauer chamber and seeded into poly-L-lysine (Sigma-Aldrich) -coated well plates with the following cell numbers: 96-well plates – 20, 000 cells in 200 µL media, 48-well plates – 40, 000 cells per well in 500 µL media, 6-well plates – 300, 000 cells in 2 ml DMEM media.

#### Determination of total receptor expression

HEK273T cells were cultured in DMEM media as described above. 6-well plates were transfected with plasmid DNA (125 ng) using lipofectamine2000 (ThermoFisher) according to the manufacturer’s protocol. A sandwich ELISA taking advantage of the HA- and Flag-Tag was used to estimate the total amounts of receptor constructs as previously described (58). In brief, after lysis of transfected HEK293T cells (10 mM

Tris/HCl, 150 mM NaCl, 1 mM DTT, 1 mM EDTA, 1% Sodium Deoxycholate, 0.2 mM NP-40) the total expression of receptors was analyzed with an anti-FLAG-M2- (Sigma Aldrich) coated to microtiter plates (10 µg/ml anti-Flag-antibody in 0.15M Sodium tetraborat/HCl, pH 8). After washing with PBS-T (0.05% Triton X-100 in in PBS) unspecific binding was blocked with DMEM + 10% FBS for 1 h at 37 °C followed by incubation of the solubilized cells for a further 1 h. Detection was achieved using an anti-HA-peroxidase-conjugated antibody (Roche) diluted 1:1, 000 in blocking solution and o-phenylenediamine (0.1 M citric acid, 0.1 M Na2HPO4) containing 0.2% H2O2. Using 1 M HCL the reaction was stopped and OD readings recorded at 492 nm and 620 nm (Tecan Sunrise).

#### Determination of cell surface expression

Cell surface expression of *AVPR2* mutants was determined in HEK273T cells using a direct ELISA (59) (Supplementary Fig. 5). In this assay, anti-HA-POD-conjugated antibody binds to the introduced N-terminal HA-tag of receptors present at the cell surface. Thereto, 24 hours after seeding into 48 well plates, cells were transfected with different amounts of plasmid DNA (25 ng) using lipofectamine2000 (ThermoFisher) according to the manufacturer’s protocol. 48 hours post transfection, cells were fixed with formaldehyde for 20 min at room temperature before blocking (DMEM + 10% FBS) for 1 h at 37°C. Later, cells were incubated with anti-HA-peroxidase-conjugated antibody (Roche) diluted 1:1, 000 in blocking solution. Detection of receptor expression on the cell surface was performed using *o*-phenylenediamine solved in substrate buffer (0.1 M citric acid, 0.1 M Na_2_HPO_4_) containing 0.2% H_2_O_2_. The reaction was stopped after 3 minutes with 1 M HCl. OD values were measured using the Sunrise microplate reader (Tecan) and normalized by detracting background (620 nm) readings at 492 nm. Expression is given after subtracting the OD values of mock-transfected cells in percentage compared to wt *AVPR2* pL.

#### Determination of receptor activity

The AVPR2 is coupled to the Gs protein and its activation results in an increase in intracellular cAMP which can be detected using the AlphaScreen^TM^ cAMP functional assay (PerkinElmer). Thus, 24 hours after seeding into 96-well plates, cells were transfected with 12.5 ng of plasmid DNA per well. 48 hours after the transfection, cells were washed with phenol red-free DMEM containing 1 mM IBMX (Sigma-Aldrich), an inhibitor of phosphodiesterases. Cells were then stimulated for 30 minutes 1 µM AVP (Sigma-Aldrich) at 37°C in media containing IBMX. To stop the stimulation, plates were placed onto ice, media was removed, and 20 µL of lysis buffer (20 mM Tween 20, 1 mM IBMX, 5 mM HEPES and BSA diluted in ddH_2_O, pH=7, 4) was added to each well. Samples were stored at -20°C until measurement. Determination of cAMP was performed according to the manufacturer’s specifications using EnVision 2105 Multimode Plate Reader (PerkinElmer).

#### Statistical Analysis

Statistical and graphical analyses were performed using Python. All experiments were performed as technical triplicates in 4 separate batches. Each tested variant is presented as % of the corresponding wild type value in the batch. Statistical significance was determined using Wilcoxon-Mann-Whitney test. P-values below 0.05 were considered to be significant.

## RESULTS

### Genes with known functional relevance are more likely to exhibit intolerance to synonymous variation

We initially hypothesized that if synonymous variation affects gene function, genes intolerant to loss-of-function or missense variation would be more likely to show depletion of sSNVs compared to neutrally evolving genes.

To test this, we used a pooled Z proportion test (synonymous Z-score) to calculate the difference in the proportion of observed and not observed sSNVs (e.g, those absent in the general population) within each gene. A high proportion of not observed variants may indicate negative selection due to functional impairment.

For this analysis, we used all human protein-coding transcripts from Ensembl Biomart (60), filtering out those with unknown nucleotides, missing start/stop codons, or patched chromosome IDs. We annotated sSNVs using synVEP v1 (22) with an updated reference from gnomAD v4 (61), classifying variants as either “observed” or “not seen”.

We then calculated Z-scores (from proportion of “not seen” to “observed” variants for a gene) for various previously reported metrics related to sSNVs: synVEP (functional effect of synonymous variants), MES (splice site strength), SpliceAI (splice site prediction), GERP++ (evolutionary conservation), RSCU and dRSCU (codon usage), CpG and CpG exon content, and distances to pre-mRNA (f_premRNA) and mRNA (f_mRNA). Only sSNVs passing synVEP quality checks were included.

Genes depleted from a specific class of variation, based on gnomAD data, where high pLI (≥ 0.8) indicates intolerance to loss-of-function and high missense Z-score (≥ 3) suggests intolerance to missense variation (61), were considered functionally constrained. We compared synonymous Z-scores between genes with high and low pLI, as well as between genes with high and low missense Z-scores.

Our analysis revealed that genes under stronger selection (high pLI or missense Z-score) exhibited significantly higher synonymous Z-scores (Figs. 1A, B) for several metrics: synVEP, MES, SpliceAI, SURF (RNA stability), GERP++, and dRSCU. However, RSCU, CpG, CpG exon, and distances to pre-mRNA and mRNA did not show significantly higher synonymous Z-scores in functionally constrained genes (Supplementary Fig. 1).

**Fig. 1.**
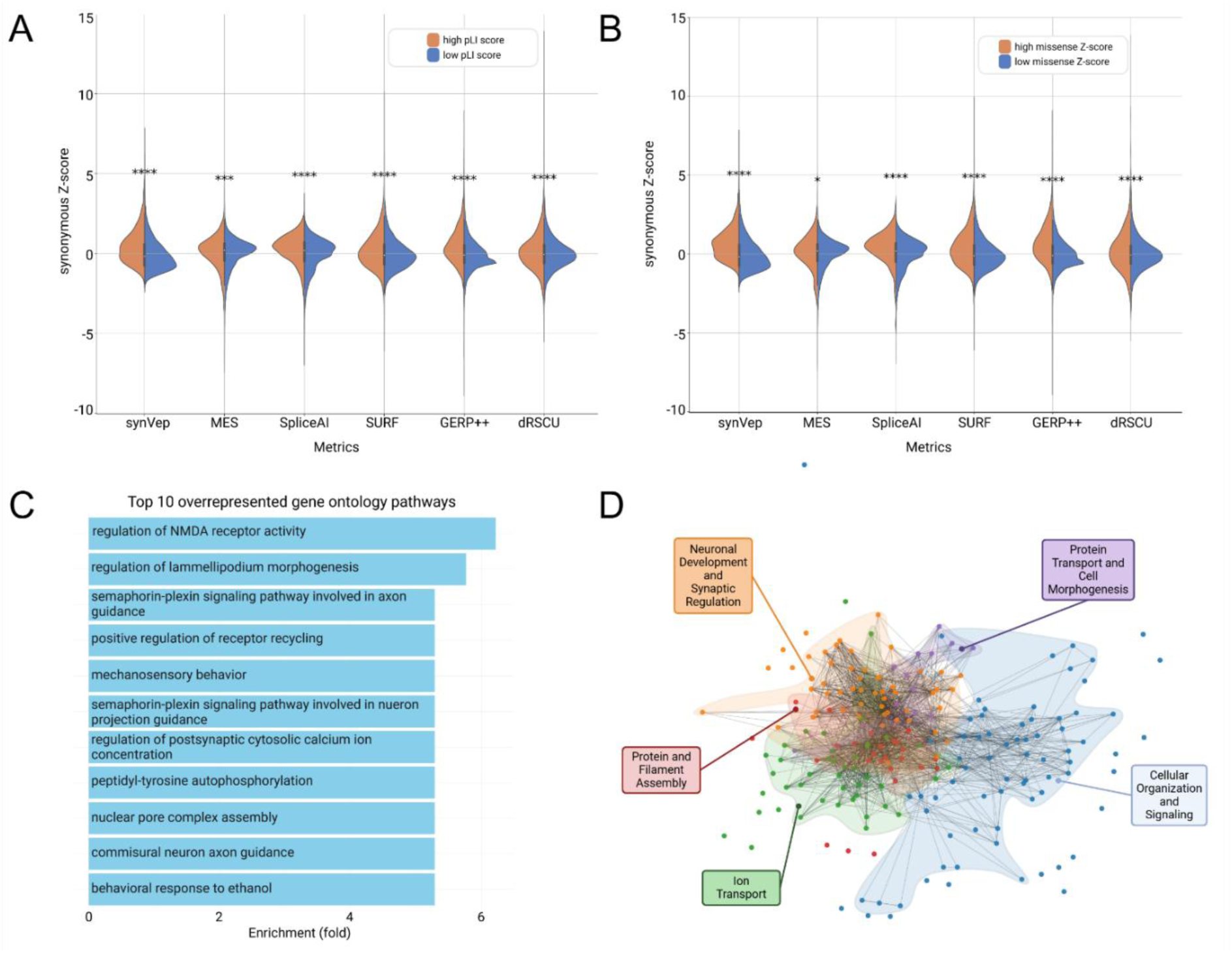
Comparison of Z-scores for synonymous variants across previously reported metrics in genes categorized by (A) high and low pLI (probability of loss-of-function intolerance) scores or by (B) high and low missense Z-scores (intolerance to missense variation). The significantly (Mann-Whitney test) higher synonymous Z-scores were observed among the metrics derived from synVep, MES, spliceAI, SURF, GERP++, and dRSCU, suggesting higher scores for genes that are intolerant to functional variation. (C) Top 10 Overrepresented gene ontology (GO) categories for genes with high synonymous Z-scores (≥ 1.96) in at least one metric. Genes with high synonymous Z-Scores show enrichment among pathways related to brain and nervous system pathways and cellular activities. (D) Clustering of GO pathways based on gene community detection. The clustering of pathways is based on the co-occurrence of genes with high synonymous Z-scores across different GO categories. The analysis reveals distinct clusters of pathways that are enriched in genes (nodes in the network) frequently found together (edges that connect nodes), which identified five key biological themes. * p-value < 0.05, *** p-value ≤ 0.001, **** p-value ≤ 0.0001.

This suggests that the high proportion of not seen sSNVs in these genes may reflect their deleteriousness, leading to negative selection. Among 3, 295 genes with high pLI, 674 had a high synonymous Z-score (≥ 1.96) across all significant metrics. Similarly, 394 of the 1, 589 genes with a high missense Z-score showed a high synonymous Z-score across all significant metrics. In total, we identified 1, 865 genes with a high synonymous Z-score (≥ 1.96) in at least one metric, regardless of their intolerance to loss-of-function or missense variation (Supplementary Table 1).

To explore whether genes intolerant to synonymous variation are enriched in specific biological processes, we conducted a gene ontology (GO) enrichment analysis. The top 10 overrepresented pathways were predominantly related to synaptic and neuronal development (Fig. 1C).

For further insight, we applied the Leiden Algorithm (62), a complex network analysis method for community detection. This approach groups genes based on their GO network connections, revealing functional relationships. We configured the analysis with a maximum of 10 iterations and a fixed random seed for reproducibility, resulting in five clusters. Most genes in these clusters were associated with nervous system function and cellular processes (Fig. 1D).

Taken together, the depletion of synonymous variation in functionally constrained genes suggests that synonymous variants may have functional effects and are subject to negative selection, as recently proposed by Gudkov et al. (17).

### *ExtraTreesClassifier* best discriminates between “benign” and “deleterious” classes in a public dataset

While deterministic approaches provide explainability, they are insufficient for reliably determining the deleteriousness of an sSNV. To achieve a more generalized and conclusive classification of sSNVs as either “benign” or “deleterious, “ we developed a supervised ensemble machine learning model. This model integrates 11 effect measures related to splicing, RNA stability, distance to premature and mature mRNA, codon usage, and CpG content, along with allele frequency in the general population, to predict the probability of deleteriousness (workflow in Fig. 2A). Our approach leverages existing public databases of known deleterious and benign genetic variants (26), including the Database of Deleterious Synonymous Mutations (dbDSM v1.2) (49) and ClinVar database (26, 50).

**Fig. 2.**
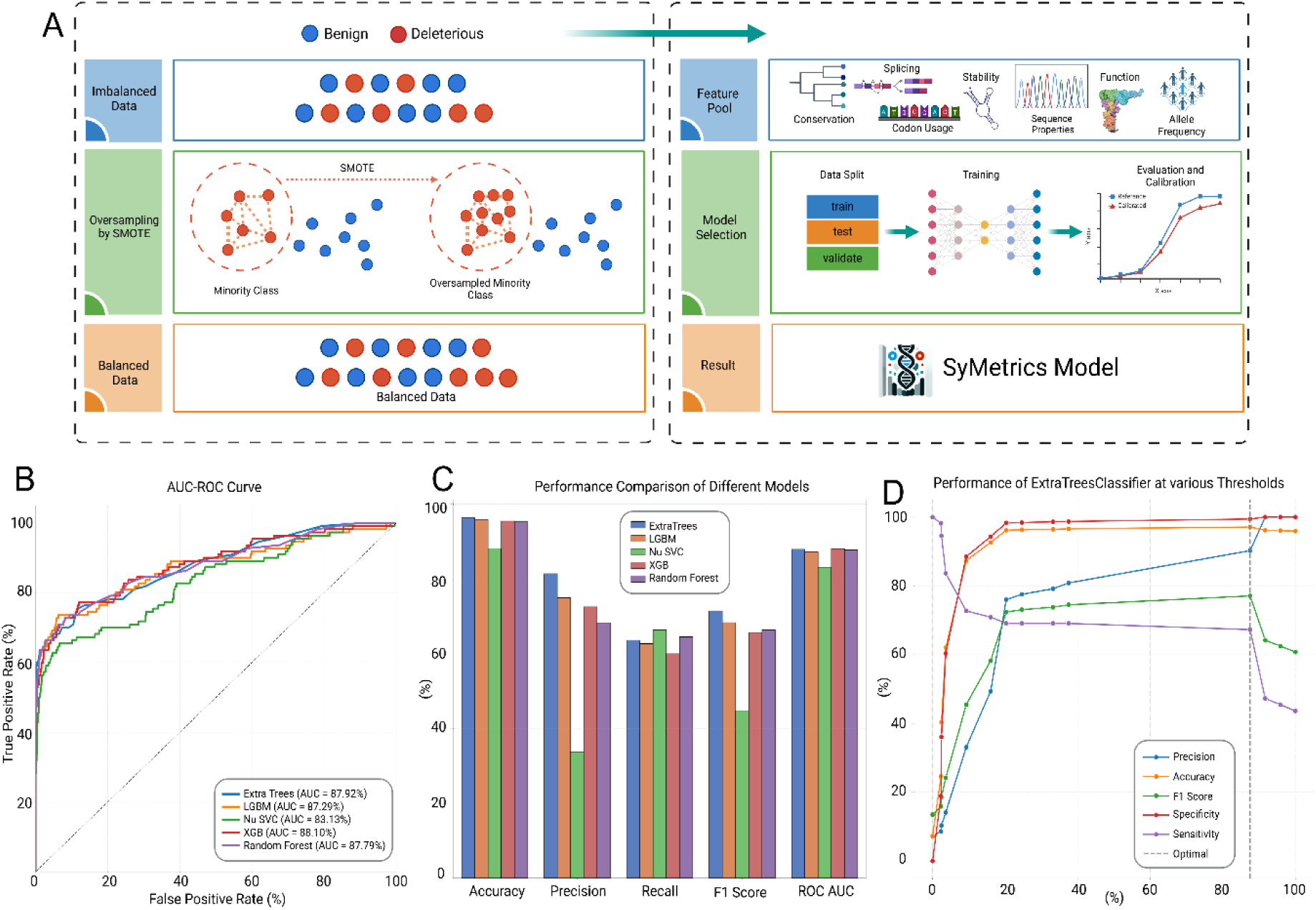
**(A) Overview of model construction.** This flowchart outlines the process of model construction, encompassing data preprocessing, training, evaluation, and calibration. During data preprocessing, an oversampling strategy was applied using SMOTE (Synthetic Minority Over-sampling Technique) to address class imbalances. The balanced dataset was then annotated with scores corresponding to selected features and subsequently divided into training, testing, and validation subsets. Before finalizing the model, we calibrated it to optimize and balance the performance metrics. **(B) Area Under the Curve (AUC) for Receiver Operating Characteristic (ROC) to evaluate model performance evaluation of top 5 models identified by Lazy Predict.** All models show good discrimination between classes (AUC > 0.8). **(C) Comparison of top 5 models identified by Lazy Predict.** The dataset consisted of 457 synonymous variants (benign: 234, deleterious: 223). The different models were used to classify the variants. The model based on *ExtraTreesClassifier* (blue) outperforms other models in most metrics (metrics definition available in Table 1). **(D) Threshold adjustment affects the performance of *ExtraTreesClassifier*.** Using the calibrated model, we determined the best threshold that can yield a good balance of all the metrics. We used the prediction probabilities and adjusted the threshold from 0 to 1 and recalculated the metrics based on each threshold. The best threshold (marked with gray vertical line) shows the point where all the metrics have high values and hence represents the optimum. (Created in BioRender. Garten, A. (2025) https://BioRender.com/ ove998a)

We incorporated 12 features - the 11 effect measures plus allele frequency - and used Lazy Predict (63) in Python to estimate the performance of different models before building them. The best-performing models in terms of accuracy, balanced accuracy, F1-score, and ROC-AUC were predominantly tree-based ensemble machine learning models (Supplementary Table 1 – Ranked models in Lazy Predict; Supplementary Fig. 2).

We then trained and evaluated the top five models predicted to perform best. Consistent with the Lazy Predict evaluation, the Extremely Randomized Trees (*ExtraTreesClassifier*) emerged as the top-performing model (Fig. 2C; Table 1). ExtraTrees is an ensemble learning technique that trains multiple decision trees and aggregates their outputs to classify variants. Unlike standard decision trees, ExtraTrees introduces additional randomness by selecting features for splitting at random, reducing bias while maintaining diversity among trees (64).

To enhance model reliability, we calibrated the trained ExtraTrees model using hyperparameters identified via Lazy Predict. We assessed model accuracy using stratified 10-fold cross-validation, yielding a mean accuracy of 96.3% and a ROC-AUC of 0.89, meaning the model correctly distinguishes between deleterious and benign variants 89% of the time.

The final calibrated model, trained on the entire dataset, achieved 97% accuracy, a ROC-AUC of 0.87, and higher precision compared to the uncalibrated version (Supplementary Fig. 3). While a default classification threshold of 0.5 is commonly used, we evaluated performance across multiple thresholds to identify an optimal balance between accuracy, precision, and recall. The most balanced performance was observed at t = 0.875, with: accuracy = 97.10%, precision = 90.23%, recall/sensitivity = 67.27%, specificity = 99.43%, and F1 Score = 0.77 (Fig. 2D). The full reference table for threshold adjustments is available in Supplementary Table 1 – Threshold Adjustment. The model output score represents the probability of a variant being deleterious, termed SyMetrics Probability.

Our analysis demonstrates that *ExtraTreesClassifier* is the most effective model for distinguishing between benign and deleterious sSNVs, achieving high accuracy and robustness through ensemble learning and calibrated threshold optimization.

### SyMetrics demonstrates a strong performance in predicting the deleteriousness of sSNVs compared to individual variant effect measures

We further evaluated the performance of SyMetrics by comparing it against individual variant effect measures - synVEP, SpliceAI, SURF, MES, and GERP++ - to assess its predictive power relative to the individual effect measures incorporated in the model. These metrics were selected because they directly relate to deleteriousness and selection. Additionally, we compared SyMetrics to CADD (65), an established and reliable score for detecting deleterious variants.

For this analysis, only variants found in dbDSM v1.2 (26, 49) were retained to represent the positive class. dbDSM v1.2 is a manually curated database of deleterious sSNVs compiled from published literature and resources such as ClinVar, GRASP, GWAS Catalog, GWASdb, PolymiRTS, PubMed, and Web of Knowledge. The negative class (neutral variants) was extracted from VariSNP (21, 66), which contains benign variants.

The variants were annotated based on their determined classes per variant effect measures using a simple deterministic scheme presented as:

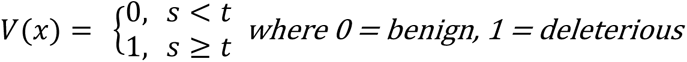

Where:

- *t* – is the set threshold for the selected effect measure
- *s* – is the effect measure score
- *V(x)* – is whether that score implies that the variant is deleterious or benign.

The thresholds selected per variant effect measures are set as follows – synVEP (t = 0.5), spliceAI (t = 0.5), SURF (t = 4), MES (t = 3), GERP++ (t = 4), and CADD (t=20).

The varying sensitivity and specificity among individual effect measures indicate that no single mechanism (e.g., splicing via SpliceAI or RNA stability via SURF) can fully capture the deleteriousness of sSNVs. However, combining multiple effect scores enhances predictive accuracy, as reflected in the SyMetrics model performance metrics: accuracy (92.69%), F1-Score (0.92), and ROC-AUC (0.93). Among the individual effect measures, SyMetrics outperformed all others, correctly identifying all 234 negative variants and 188 of the 223 positive variants (Fig. 3).

**Fig. 3.**
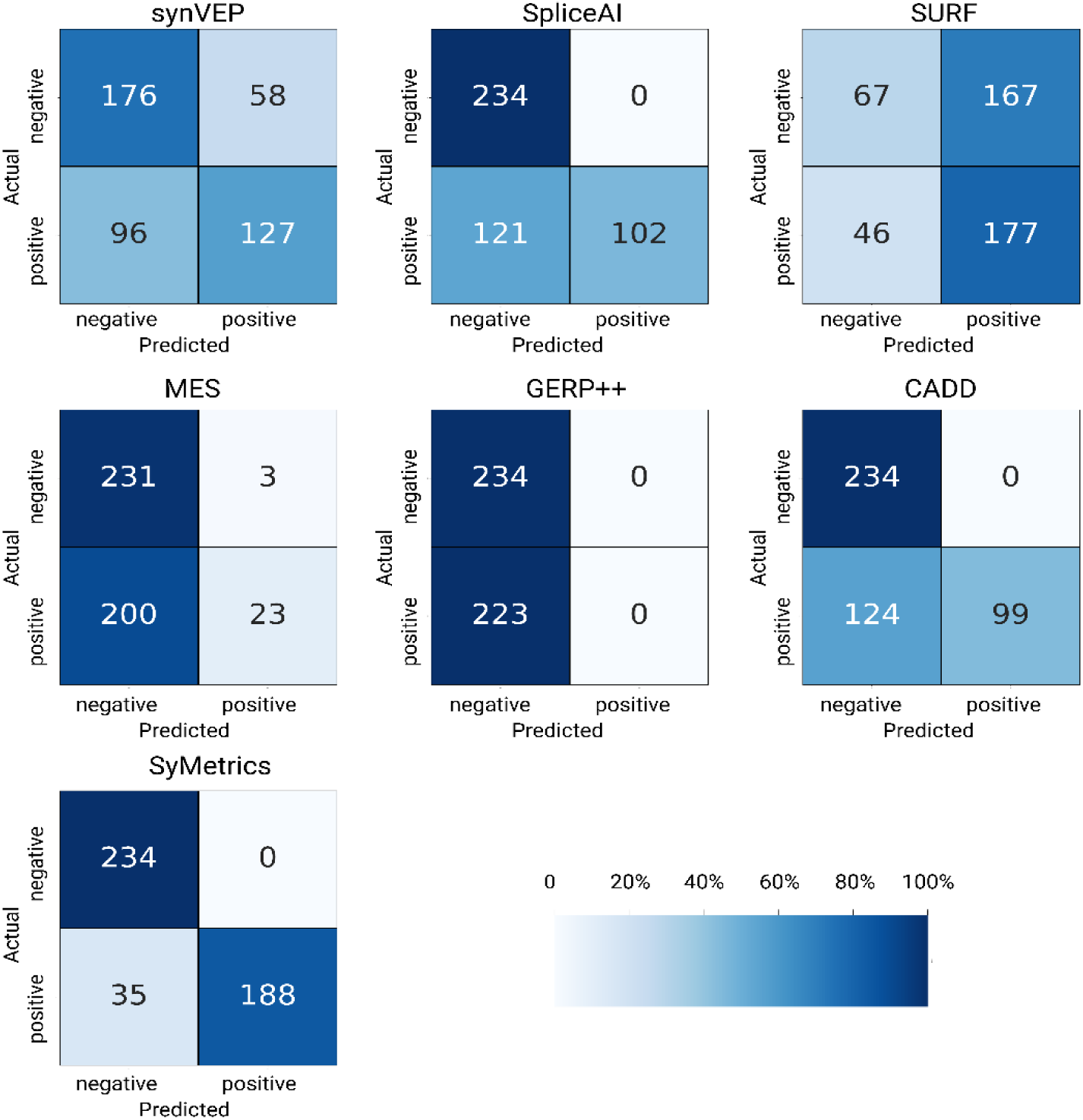
Heatmap of confusion matrices for variant effect predictions based on different metrics. The test dataset consisted of 457 synonymous variants (benign: 234, deleterious: 223). We calculated the number of true positive, false positives, true negatives, and false negatives, depicted across actual and predicted, for each metric based on the recommended thresholds. Compared to all other metrics SyMetrics performed best in classifying the actual/true class among others.

In sum, SyMetrics outperforms individual variant effect measures in predicting the deleteriousness of sSNVs, achieving higher accuracy and reliability by integrating multiple predictive features.

### Predicted Deleterious sSNVs are depleted in general population and enriched in genes under strong selection

To determine whether predicted deleterious sSNVs are depleted in the general population, we scored the generated sSNVs and categorized them by allele frequency (AF) into five groups:

very common, common, low frequency, rare, and not seen. We then performed a pairwise comparison of the number of sSNVs with a SyMetrics score exceeding the threshold (t = 0.875) across the following groups: common (0.05 < AF < 0.1) vs. very common (0.1 < AF < 1), common vs. low-frequency variants (0.01 < AF < 0.05), common vs. rare (0 < AF < 0.01), and common vs. not seen (AF = 0).

We observed significant differences in score distribution for all pairwise comparisons except for the very common group. This suggests that variants with higher SyMetrics scores are depleted in the general population due to selection.

To further investigate these findings, we calculated the odds ratio of a variant being deleterious in ‘very common, low-frequency, rare, and not seen’ groups relative to the ‘common’ group using Fisher’s Exact Test. The predicted deleterious sSNVs were found to be significantly enriched among low-frequency, rare, and not seen groups (Fig. 4A).

**Fig. 4.**
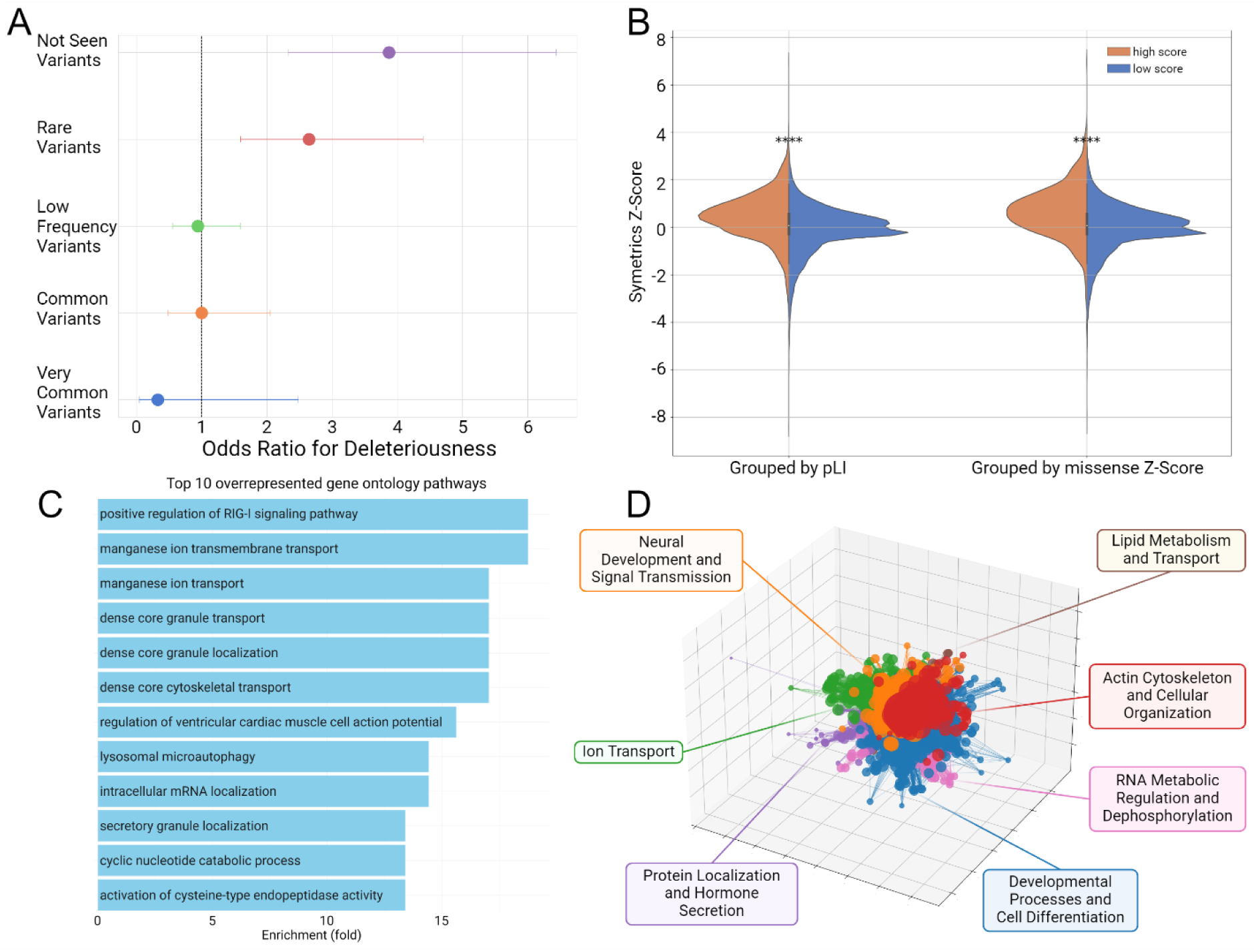
**(A) Odds ratios for deleteriousness based on SyMetrics prediction across synonymous variants with different allele frequencies in general population.** Not Seen Variants = allele frequency is AF=0 in gnomAD v4, Rare Variants = allele frequency is 0 < AF < 0.01 in gnomAD v4, Low Frequency Variants = allele frequency is 0.01 < AF < 0.05 in gnomAD v4, Common Variants = allele frequency is AF > 0.05 in gnomAD v4, Very Common Variants = allele frequency is 0.1 < AF < 1 in gnomAD v4. **(B) Comparison of synonymous Z-Scores for genes with high versus low pLI and missense Z-Scores.** Genes with high pLI and missense Z-Score have significantly higher synonymous Z-scores **** p-value ≤ 0.0001, Mann-Whitney test. **(C) Top 10 gene ontology categories enriched in genes with high synonymous Z-score.** Using the Mann-Whitney test, we test whether the significant synonymous Z scores of genes with high missense z score is higher than the one with low missense z scores. The significantly higher SyMetrics Z-scores were derived from the SyMetrics Probability. **(C) Top 10 Overrepresented gene ontology (GO) categories for genes with high SyMetrics Z-scores (≥ 1.96). (D) Clustering of GO pathways based on gene community detection.** The clustering of pathways is based on the co-occurrence of genes with high synonymous Z-scores across different GO categories. The analysis reveals distinct clusters of pathways that are enriched in genes (nodes in the network) frequently found together (edges that connect nodes), which identified seven key biological themes, among which developmental processes and cell differentiation is the best represented.

Using stratified bootstrapping, we estimated the proportion of potentially deleterious synonymous variants: 1.98 ± 0.15% in the not seen category, 1.60 ± 0.15% in singletons, 1.07 ± 0.13% in rare variants, 0.53 ± 0.1% in common variants. This corresponds to n = 931, 841 (95% CI: 861, 247 – 1, 002, 435) deleterious variants in the not seen group.

Next, we compared synonymous Z-scores using SyMetrics Probability under the same grouping scheme, further stratifying genes by their pLI (probability of loss-of-function intolerance) and missense Z-score. As expected, genes under stronger selection (high pLI ≥ 0.8 or missense Z-score ≥ 3) exhibited significantly higher synonymous Z-Scores (Fig. 4B), reinforcing the idea that sSNVs in these genes are absent in the general population due to their deleteriousness.

Altogether, we identified 309 genes with a high synonymous Z-score (> 1.96), independent of the grouping scheme based on pLI or missense Z-score (Supplementary Table 1 – High SyMetrics Z). In sum, predicted deleterious sSNVs are significantly depleted in the general population and enriched in genes under strong selection, highlighting their potential functional impact.

### Experimental validation of sSNVs affecting *AVPR2* function

To determine the extent to which our predictions can be validated, we conducted functional testing of eight variants in the *AVPR2* gene. *AVPR2* encodes the arginine vasopressin receptor type 2, and pathogenic variants in this gene cause X-linked nephrogenic diabetes insipidus type 1. To mitigate potential bias toward functionally critical genes, we selected an X chromosome-located gene for which no constraint metrics are currently available in gnomAD v4. Additionally, we chose *AVPR2* because it does not exhibit an overall synonymous Z-score indicative of intolerance to synonymous variation (Z-score: 1.006).

None of the predicted deleterious variants was observed in the general population (gnomAD v4). The variants are distributed across exons 2 and 3. Additionally, we included two predicted benign variants in our analysis.

We evaluated the impact of these variants on receptor function by measuring total cellular and cell surface expression (indicators of protein production, folding, and/or trafficking defects) and cAMP formation (agonist-induced receptor activation) in comparison with the wild-type receptor. We used minigene constructs containing all three exons, interrupted by their natural introns, cloned into a eukaryotic expression vector.

Among the investigated variants, three had a SpliceAI score >0.5 (Supplementary Table 1 – *AVPR2* Variants). Interestingly, the variant NM_000054.7:c.27T>A, p.(Ala9=), with a SpliceAI score of 0.99 and SyMetrics 0.875, showed no functional effect (Figs. 5A, B, C). However, variant 2 (NM_000054.7:c.276A>G, p.(Gln92=)) exhibited the lowest expression and the strongest functional effect among all tested variants (Fig. 5). While the SpliceAI score of this variant is 0.5, the SURF score is 27.54, suggesting high mRNA instability.

**Fig. 5.**
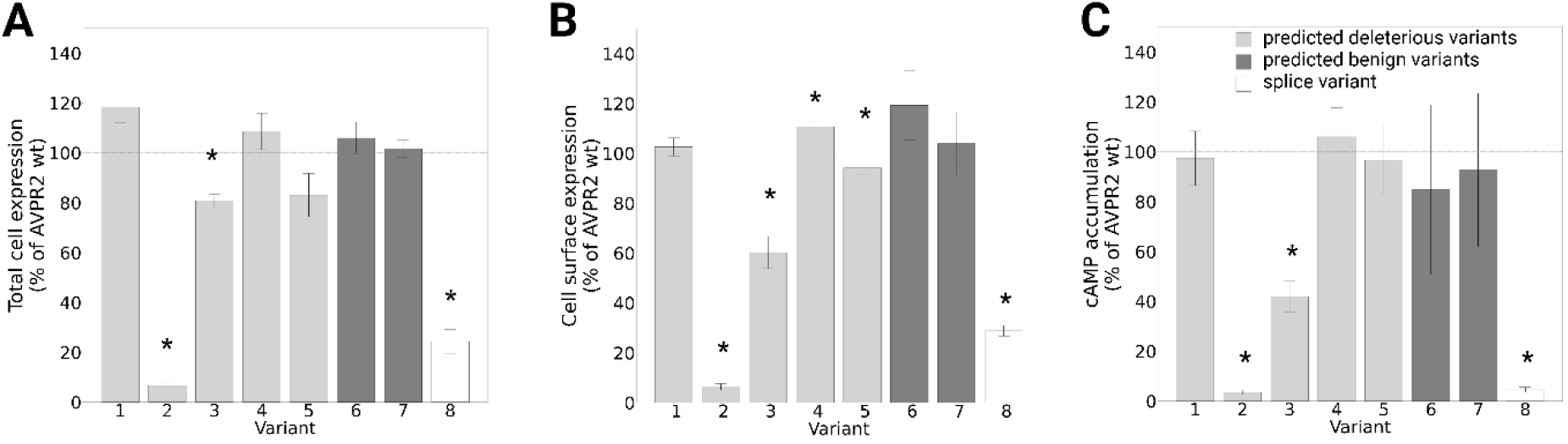
Synonymous variants affect the functionality of the human vasopressin 2 receptor (encoded by *AVPR2* gene). We predicted sSNVs that do not alter the amino acid residue but may affect receptor functions — either predicted to be deleterious (light grey) or benign (dark grey). These variants were inserted into the genomic sequence of the human *AVPR2* and functionally tested them **(A)** First, total receptor protein expression was determined in an ELISA taking advantage of the N-terminal HA- and the C-terminal Flag-tag. **(B)** Then, cell surface expression of the receptor protein was determined. **(C)** Finally, cAMP accumulation was detected after receptor stimulation. For all variants experiments show a consistent trend, albeit only for variants 2, 3, and 8 all experiments showed a significant result. * p-value (Wilcoxon-Mann-Whitney test) < 0.05. All experiments are presented as a percentage of the corresponding matched wild-type receptor. Dashed horizontal line represents wild-type level (100 %).

To assess whether a similar level of protein reduction could be observed for another splicing variant, we introduced a variant in the canonical splice site (NM_000054.7:c.910+2T>C, p.?, variant 8). This mutation is expected to result in the loss of an acceptor site, intron retention of 20 base pairs, and a premature stop codon. While the reduction in protein expression was significant, it was less pronounced than that observed for variant 2.

For variant 3, we also identified reduced cell surface expression, although to a lesser extent than in variant 2. Splicing prediction indicates a donor site loss (SpliceAI = 0.62), leading to the same transcript as the canonical splice site variant. However, variant 3 exhibited significantly higher expression than the canonical splice site variant.

Lastly, we examined two predicted deleterious variants: NM_000054.7:c.948C>A, p.(Leu316=) (variant 4) and NM_000054.7:c.960C>T, p.(Thr320=) (variant 5), with SpliceAI scores of 0.23 and 0.42, respectively, suggesting a low probability of having a splicing effect. However, both had SURF scores exceeding the significance threshold of 5, suggesting potential effects on RNA stability. Interestingly, variant 4 exhibited significantly higher cell surface expression (Fig. 5B), while variant 5 showed significantly lower cell surface expression. However, total cell expression for these two variants did not reach significance, although the trend followed the same direction (Fig. 5). A similar pattern was observed in cAMP accumulation, but due to high variability, it did not reach statistical significance. We also tested two variants predicted to be benign (variants 6 and 7), both of which showed no significant difference from the wild type. In conclusion, among the five predicted deleterious variants, we observed significant alterations in cell surface expression in four cases, including one with higher expression than the wild type. Considering RNA stability may improve predictions of a variant’s deleteriousness, particularly in cases of potential gain-of-function effects, such as variant 4.

### SyMetrics predicts deleterious potential of sSNVs identified in a patient cohort

At the Human Genetics Institute at Leipzig University Clinics, we reanalyzed data from 11, 903 patients who had undergone genetic evaluation either through exome/trio-exome sequencing or using a morbid gene panel (TruSight One, Illumina). We identified 767, 774 unique variants, which were scored using SyMetrics. Of these, 8, 088 sSNVs were predicted to be deleterious. To refine our analysis, we applied additional filters, requiring that the variant be absent from the general population and that the Human Phenotype Ontology (HPO) terms associated with the affected gene match those assigned to the patient, according to the HPOsim score (67). We identified 15 variants in genes that matched the patient’s phenotype and were predicted to be deleterious. Among these, nine variants were classified and reported as either likely pathogenic (n = 4) or pathogenic (n = 5). Additionally, six variants were reported as variants of uncertain significance (VUS). Parental segregation analysis was available for only three of these cases.

In one case, the variant c.3015G>A, p.(Val1005=) in *IQSEC2* occurred *de novo*. Of all the effect measures considered, only synVEP was above the deleteriousness threshold. Notably, the variant had a SpliceAI score of only 0.47, meaning it could have been missed in routine clinical screening. Unfortunately, RNA analysis was not available for this proband.

In another case, the variant c.1077G>A, p.(Lys359=) in *ARHGEF9* was hemizygous in the affected proband and inherited from a carrier mother. Since developmental and epileptic encephalopathy 8 follows X-linked inheritance, segregation analysis alone could not clarify its pathogenicity. However, we recommended further familial segregation analysis, as a maternal uncle exhibited a similar phenotype. In this case, the SpliceAI score was only 0.18, meaning the variant would likely have been missed by routine clinical screening. However, both synVEP and SURF were above the threshold, suggesting potential RNA instability.

Similarly, we identified a patient who was a homozygous carrier of the variant c.729G>A, p.(Thr243=) in *SLC39A4*, a gene associated with acrodermatitis enteropathica. Both parents were carriers, making segregation analysis uninformative. However, the variant had a SpliceAI score of 0.76, strongly suggesting a splicing effect, for which we recommended RNA analysis. In conclusion, applying SyMetrics in a clinical setting can be particularly beneficial for detecting deleterious variants that may not affect splicing and where segregation analysis can provide additional insights.

## DISCUSSION

Recent developments in sequencing technologies have greatly improved our understanding of how genetic variation is associated with phenotypes or diseases. The role of synonymous variants in the context of human disease has garnered increasing attention in recent years, challenging the traditional view that these variants are functionally neutral (15). Synonymous single nucleotide variants (sSNVs), defined as single-nucleotide changes that do not alter the amino acid sequence of proteins, can nonetheless have significant biological implications. Evidence suggests that these variants can affect gene expression by interfering with mRNA splicing, stability, and translation efficiency, potentially contributing to disease susceptibility and phenotypic variation.

The scarcity of reliable genetic datasets on synonymous variants is evident in the comparatively lower level of research and investigation they receive, especially when compared to missense or nonsense variants. However, the association of synonymous variants with various rare diseases, as well as with cancer, where 6–8% of pathogenic single nucleotide substitutions are synonymous variants, underscores the importance of studying these variants more comprehensively. These variants frequently act as driver mutations in human cancers.

The evaluation of synonymous single nucleotide variants (sSNVs) has been significantly advanced by the development of *in silico* scores that assess their potential impact on splicing, RNA stability, sequence conservation, translation efficiency, and other functional mechanisms. To explore whether genes known to be intolerant to loss-of-function or missense variation, which are often associated with disease, are also sensitive to synonymous changes, we compared the distribution of various functional scores between genes under strong selective constraints and those without such constraints. Specifically, we analyzed metrics assessing overall functional impact (synVEP), splicing effects (MES, spliceAI), RNA stability and folding (SURF), evolutionary conservation (GERP++), codon usage (RSCU, dRSCU), and sequence properties like CpG, CpG exons, and proximity to mRNA and pre-mRNA boundaries. Our analysis revealed a significantly higher distribution of five of these scores, related to splicing and RNA stability, for intolerant genes (Fig. 1). This is in line with accumulating evidence indicating that synonymous changes can modulate the stability of mRNA and its translation kinetics, ultimately affecting protein synthesis. Similarly, Gaither and colleagues showed that sSNVs that may disrupt mRNA structure have significantly lower rates in human populations

(68). Strong evidence that sSNVs can alter protein function and expression is provided by functional studies, which, although limited to particular genes, provide proof-of-concept. To this end, synonymous variants have been shown to influence blood group expression (69) or even cause hemophilia (c.459G>A (Val=) in the *F9* gene) by slowing factor IX translation and affecting its conformation, which results in decreased extracellular protein levels (70).

To assess which genes appeared to be most intolerant to synonymous variation, we performed a gene ontology analysis, which revealed clustering of these genes in neuronal development and structural cellular processes (Fig. 1C, D). This further strengthens the idea that sSNVs in these genes could have significant functional consequences, and as these are fundamental processes under strong evolutionary constraint, any disruption could have detrimental effects on the organism.

While the above-mentioned individual scores are valuable for identifying variants that disrupt specific processes, we developed a comprehensive tool that predicts the overall deleteriousness of an sSNV by integrating multiple metrics. The allele frequency in the general population of the variants appears to be an important predictor of deleteriousness, as suggested by Gaither *et al.* and Zeng *et al.* (22, 68). This is supported by our analysis, which shows a depletion of sSNVs in genes under constraint. Thus, in our model, we also integrated allele frequency in the general population (gnomAD v4) (61) as one of the parameters. We identified *ExtraTreesClassifier* to be the best-performing machine learning model, which, after calibration and setting the threshold to t = 0.875, reached an accuracy of 97.10%, a precision of 90.23%, and specificity of 99.43% (Fig. 2, Table 1). Our model, which integrates multiple aggregated metrics, demonstrated improved accuracy in identifying both benign sSNVs and those with potential pathogenic consequences (Fig. 3), outperforming individual metrics as well as the widely established CADD score (65). Interestingly, evolutionary conservation represented by GERP++ discerned all the benign variants but missed all known pathogenic sSNVs, suggesting that evolutionary conservation is a poor metric for synonymous variants. Conversely, SURF, which predicts RNA stability, managed to detect many of the true positives (177/223) but showed low specificity in discriminating against the true negatives (67/234) (Fig. 3). SpliceAI was able to properly detect all true negatives but showed lower performance for the true positives (102/223). This may suggest that many of the sSNVs may have a functional consequence other than splicing. Overall, SyMetrics performed best, properly identifying all true negatives and 188/223 (approximately 84%) of the true positives (Fig. 3).

We were then prompted to determine which genes are depleted of synonymous variation based on the SyMetrics predictions. We initially showed that variants not present in the general population have an odds ratio of approximately 4 to be deleterious (Fig. 4A). Furthermore, we reproduced the synonymous constraint, as determined by the SyMetrics Z-score for the functionally relevant genes (Fig. 4B), and finally, we identified 309 genes with a high SyMetrics Z-score (Supplementary Table 1 – High SyMetrics Z). Gene ontology clustering revealed seven key biological themes, among which developmental processes and cell differentiation were the best represented, followed by neuronal development and cytoskeletal and cellular organization. The depletion of such pathways from synonymous variation further underscores the functional relevance of sSNVs.

To confirm the functional importance of sSNVs, we tested predicted deleterious and benign variants in the *AVPR2* gene. The variant with the strongest effect was variant 2, NM_000054.7: c.276A>G, p.(Gln92=) (Fig. 5). This variant had been previously reported in patients with diabetes insipidus. It was shown that it introduces an alternative acceptor site leading to a 5’ truncated exon 2 lacking its first 251 bases (71). The truncation causes a frameshift followed by a premature stop codon, which may trigger nonsense-mediated decay. However, compared to variant 8 located in the canonical splice site, which also leads to a premature stop codon, the reduction in protein expression for variant 2 was significantly lower. This may suggest that for variant 2, RNA instability may additionally contribute to the almost absent expression of the protein, given the SURF score of 27.54. Further, for variant 3, the splicing prediction is of a donor loss (spliceAI=0.62), which would lead to the same transcript as in the case of variant 8 in the canonical splice site. However, in the case of variant 3, we observe much higher expression than in the case of variant 8. While in our functional setup we cannot test whether splicing is affected, the higher expression level of this variant makes it very likely that splicing is not affected in this case. In turn, the reduced expression could be a result of altered RNA stability. Two of the tested variants did not have high spliceAI scores. For these variants, we showed significantly altered cell surface expression (Fig. 5B). Variant 4 showed higher expression, and the SURF score in this case was 5.93. Increased mRNA stability after synonymous variation had been demonstrated for the synonymous variant in the prothrombin gene, *F2* (NM_000506.4: c.1824C>T; p.Arg608=, SURF=15.88). The variant causes increased prothrombin mRNA and plasma protein levels, such that carriers of the variant are at increased risk of thromboembolism (72). Thus, considering RNA stability may improve the prediction for deleteriousness, especially in cases where the mechanism may be gain-of-function. Although the effect of variants 4 and 5 seems more limited, the consistent trend across all tests - total expression, cell surface expression, and cAMP accumulation (Fig. 5) - strongly suggests a functional impact.

Lastly, to check whether SyMetrics could be useful in a clinical setup, we revisited 11, 903 cases presenting the suspicion of a genetic disease. We identified 15 variants that were predicted to be deleterious, and the affected gene matched the proband’s phenotype. Nine of these variants were classified according to American College of Medical Genetics (ACMG) criteria (73) as pathogenic or likely pathogenic. For the rest, we had parental information only in three cases. One of the variants in c.3015G>A, p.(Val1005=) in *IQSEC2* occurred *de novo*, making it very likely to be causative for the reported symptoms. Furthermore, we identified a hemizygous variant c.1077G>A, p.(Lys359=) in *ARHGEF9* in a proband with epileptic encephalopathy and ataxia. The variant is inherited from a carrier mother, and there is a similarly affected maternal uncle, for whom no genetic testing is available. However, the phenotype match to OMIM #300607, as well as the plausibility of the inheritance mode with multiple individuals affected in the family, makes it very likely that this is the cause of the symptoms, although the variant must still remain classified as variant of unclear significance according to ACMG. In both cases, we recommended RNA analysis, which could shed more light on the functional mechanism.

In conclusion, this study underscores the significant impact of sSNVs on essential biological processes, highlighting their potential role in disease mechanisms. The observed link between genetic variants and functional outcomes, especially in early-onset conditions like epileptic encephalopathy and related disorders, emphasizes the need to consider these variants in clinical research and diagnostics. Overlooking the functional relevance of sSNVs may lead to missed disease etiologies, ultimately limiting our understanding of the genetic foundations of complex disorders. To aid in this effort, we present a tool that predicts functionally relevant synonymous variations. This tool is freely available online and provides researchers with insights into the underlying mechanisms of a variant, displaying all relevant metrics and allele frequencies in the general population. As the availability of genetic data increases, we will likely better understand the “silence” of synonymous variants.

## ACKNOWLEDGEMENTS

We thank the German Research Foundation and University of Leipzig for the opportunity for entrusting the grants to support the projects in Le Duc Lab. The graphical abstract was created in BioRender. Garten, A. (2025) https://BioRender.com/6e69nny

## AUTHOR CONTRIBUTIONS

L.B., conceptualization; writing– original draft; formal analysis; investigation; methodology. M.S., D.T., A.S., and J.H, conceptualization, writing – review & editing; investigation; resources; methodology; validation. F.G., software; D.L.D., investigation; methodology. T.S., conceptualization, investigation, validation, writing – review & editing. S.H. – software, writing – review & editing, U.S. – software & resources. S.S., B.S., P.M., F.B., A.G. and J.K., writing – review & editing. D.L.D., conceptualization; writing– original draft; supervision; funding acquisition.

## SUPPLEMENTARY DATA

Supplementary Data is available online

## DATA AVAILABILITY

The processed files and databases used in this study can be found in Zenodo repository https://doi.org/10.5281/zenodo.11082720. The Python package can be found in PyPi repository, https://pypi.org/project/symetrics/. The web application is accessible through https://tools.hornlab.org/symetrics/

## Supplementary Figures

**Supplementary Fig. 1.**
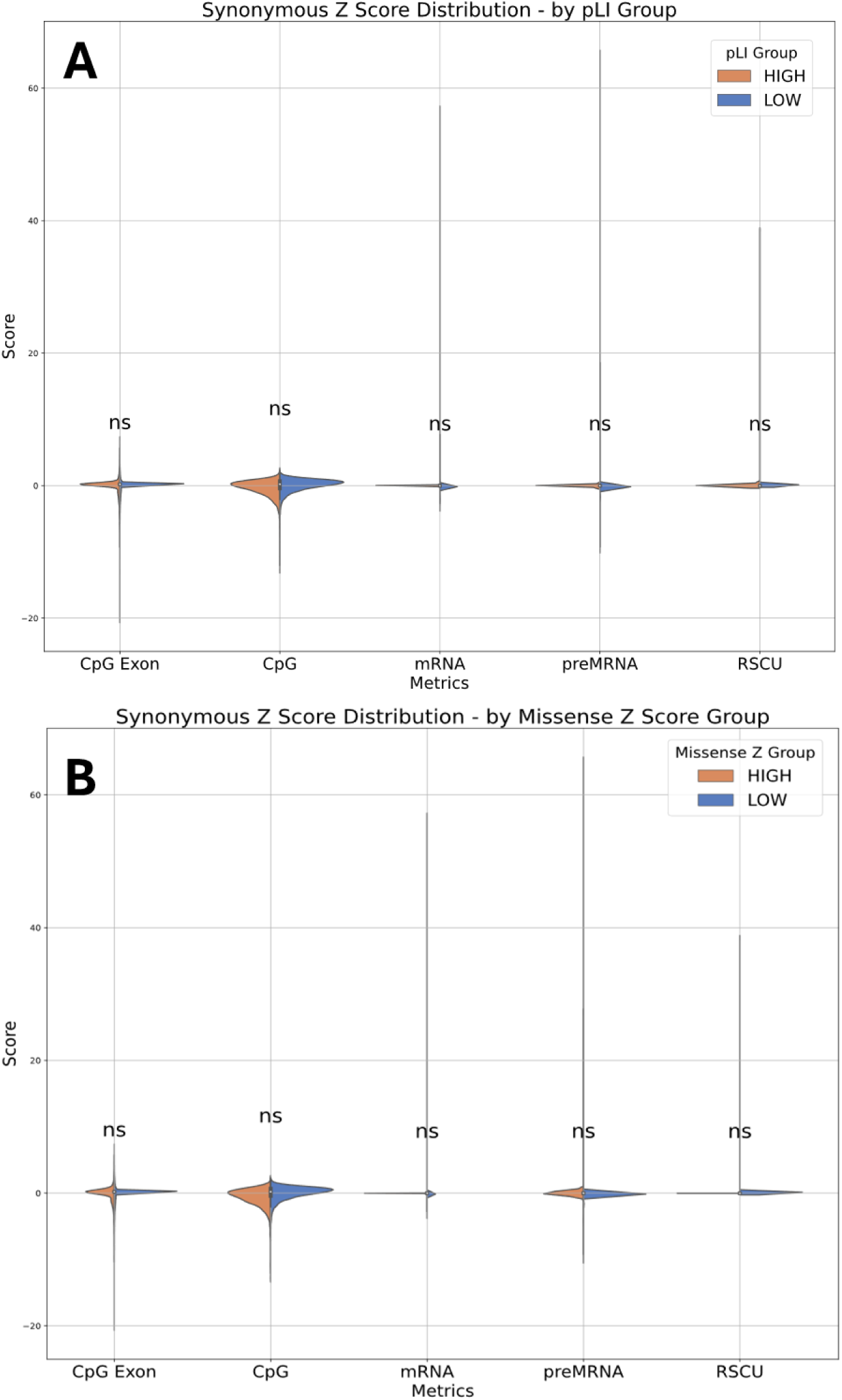
Comparison of Z-scores for synonymous variants across metrics w/o significance in genes categorized by (A) high and low pLI (probability of loss-of-function intolerance) scores or by (B) high and low missense Z-scores (intolerance to missense variation). There were no significantly higher proportion, or higher synonymous Z-scores observed among the metrics using the CpG Exon, CpG, distance to pre-mRNA and mRNA, and RSCU suggesting tolerance to variation related to mechanisms reflected by those metrics.

**Supplementary Fig. 2.**
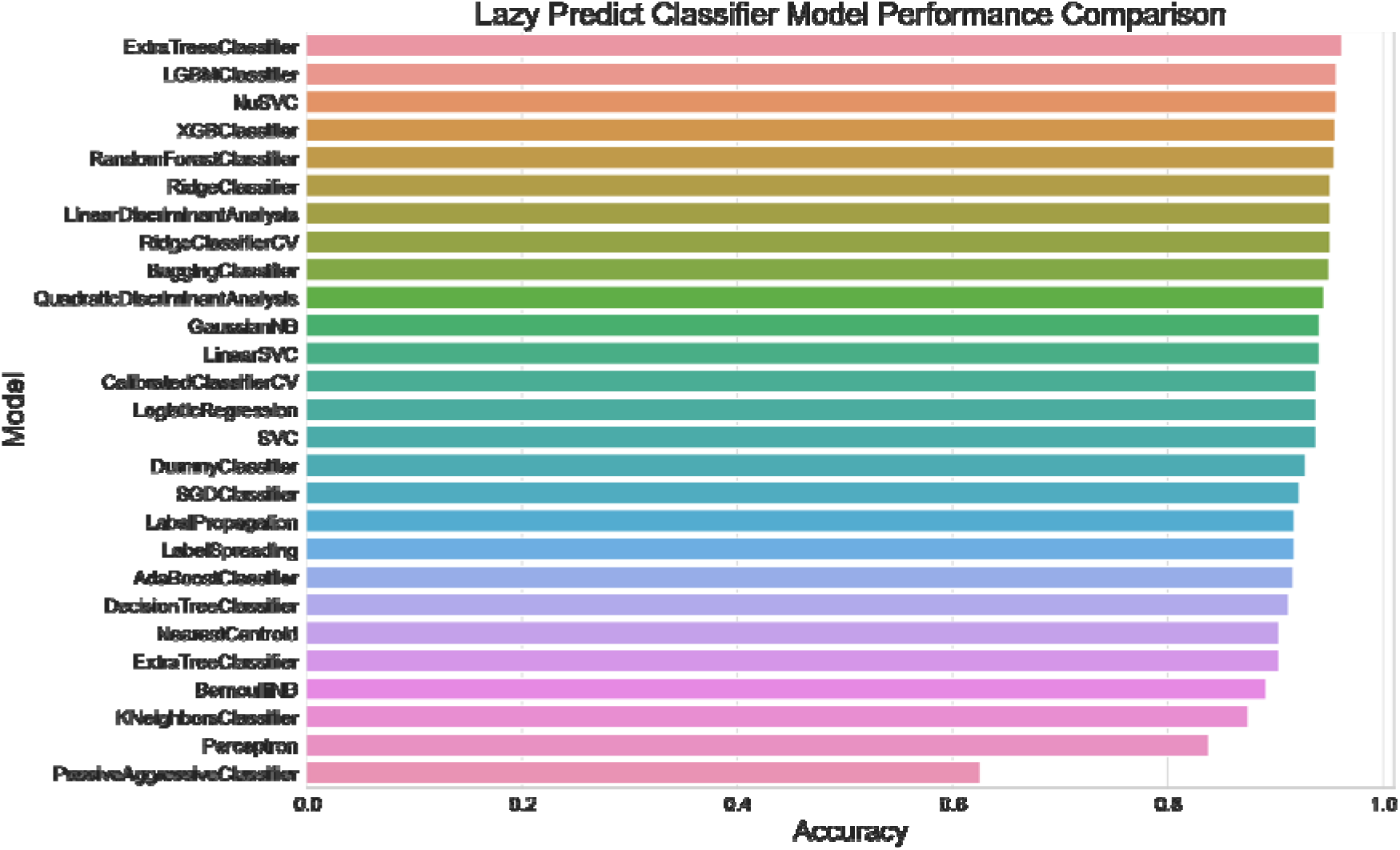
Comparison of all models in LazyPredict. Using LazyPredict, we estimated the performance of all possible classification models within the framework and ranked them by selected metrics – in this case accuracy. The results show that ensemble-based methods such as *ExtraTreesClassifier*, *LGBMClassifier*, and *XGBClassifier* dominate the top position. We proceeded with the top 5 for further comparison and selection of model.

**Supplementary Fig. 3.**
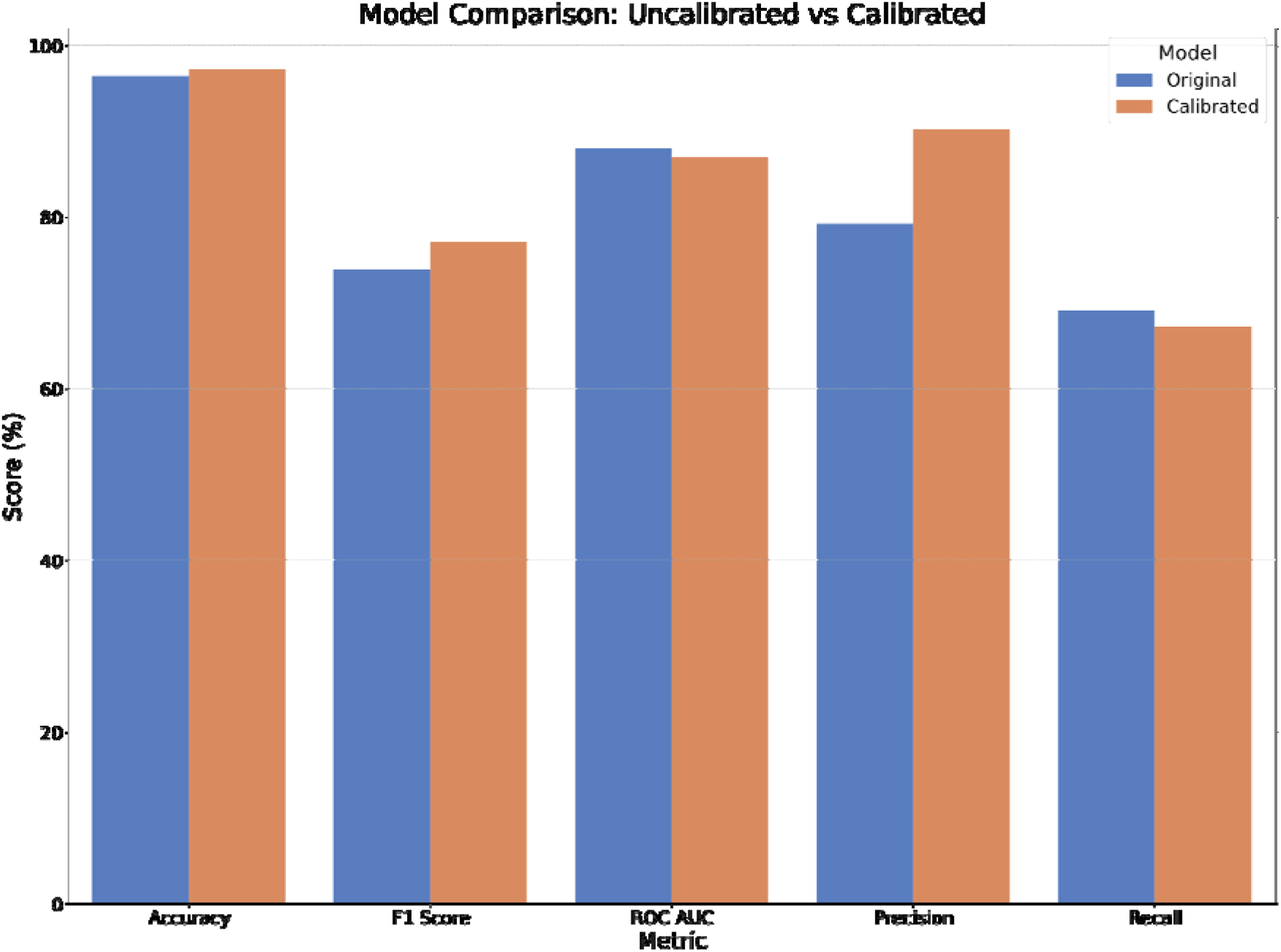
Comparison of uncalibrated and calibrated *ExtraTreesClassifier*. This analysis evaluates the performance improvement after model calibration, where the prefit *ExtraTreesClassifier* is followed by Isotonic Regression to adjust output probabilities to reflect true likelihood. While overall accuracy shows a slight increase, both F1-score and precision improve substantially, demonstrating that calibrated models enhance specific performance aspects. Trade-offs between performance metrics should be considered in the decision for calibration.

**Supplementary Fig. 4.**
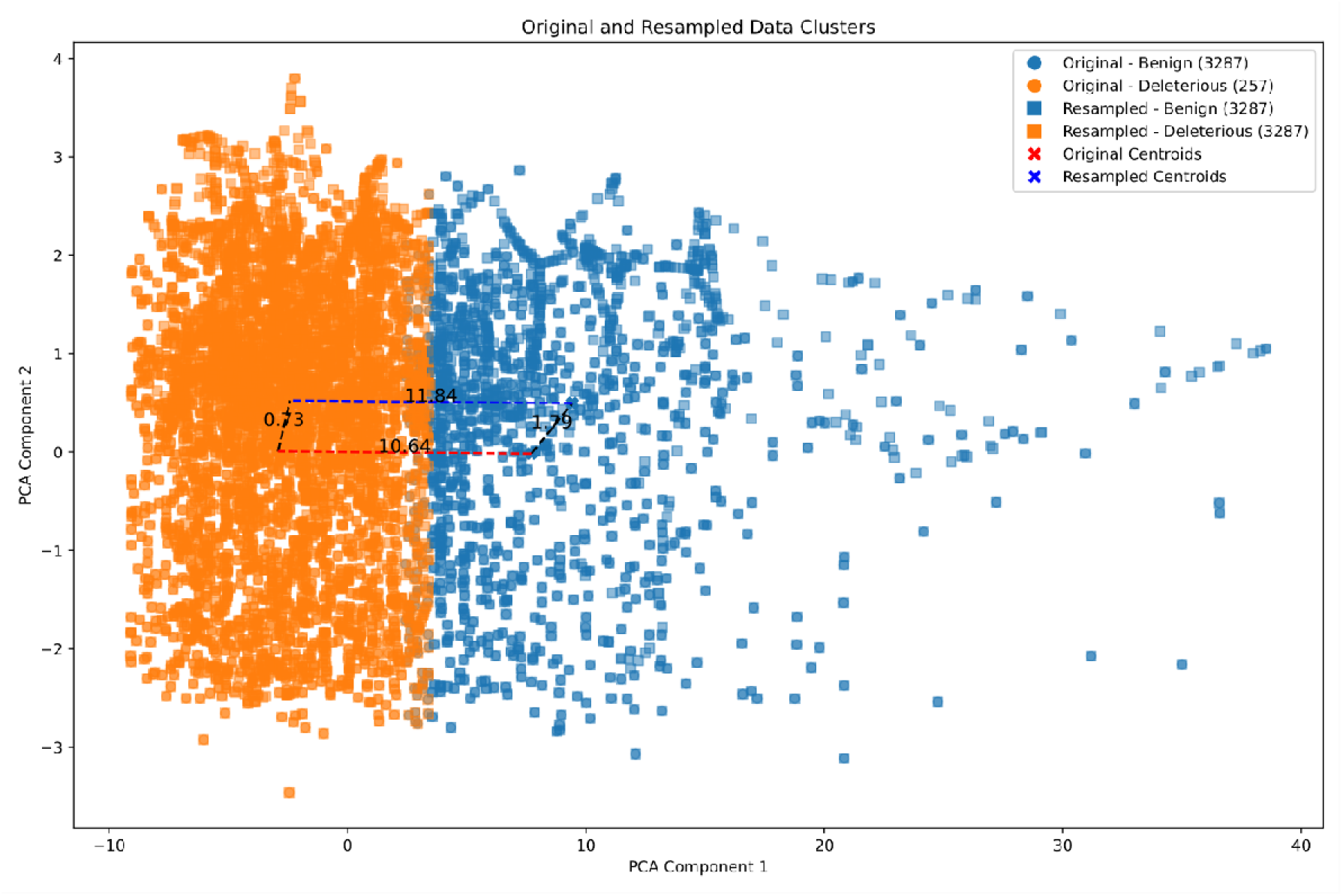
Clustering of “benign” and “deleterious” classes before and after oversampling. The „deleterious” class was oversampled using SMOTE to increase its representation. To ensure that data quality and integrity were maintained, we compared the clustering of the classes before and after oversampling. The centroid distance between the original and resampled deleterious clusters (*d = 0.73*) is much smaller than the interclass distance between deleterious and benign clusters (*d > 10*), indicating that the behavior and properties of the original and resampled data are similar.

**Supplementary Fig. 5.**
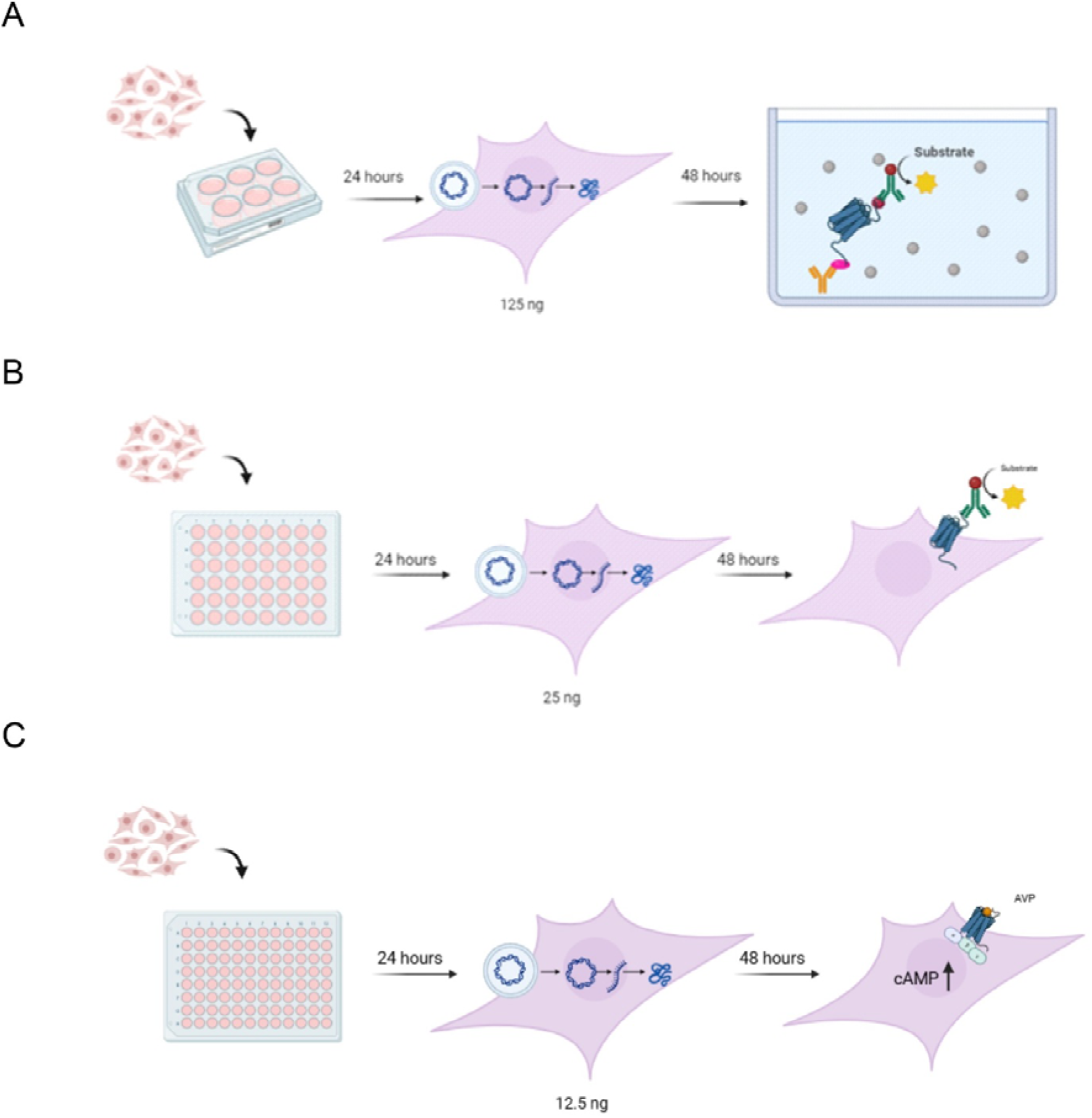
Overview of functional experiments to determine the impact of *AVPR2* mutants. **(A)** To analyze total receptor protein, cells were seeded into 6 well plates and transfected with 125 ng plasmid DNA encoding either wild type (wt) receptor or the indicated mutants. Using a sandwich ELISA approach the amount of total receptor was determined relative to wt AVPR2 expression. **(B)** Expression on the cell surface was determined in 48 well plates with cells transfected with 25 ng receptor-encoding plasmid. Detection was performed using a direct ELISA with a POD-coupled antibody against the N-terminal HA-tag. **(C)** To assess receptor activity, the amount of intracellular cAMP after stimulation with the agonist AVP was determined.

## REFERENCES

1. Sauna, Z.E. and Kimchi-Sarfaty, C. (2011) Understanding the contribution of synonymous mutations to human disease. Nat Rev Genet, 12, 683–91.

2. Winterer, G. and Goldman, D. (2003) Genetics of human prefrontal function. Brain Res Brain Res Rev, 43, 134–63.

3. Hunt, R.C., Simhadri, V.L., Iandoli, M., Sauna, Z.E. and Kimchi-Sarfaty, C. (2014) Exposing synonymous mutations. Trends Genet, 30, 308–21.

4. Jaganathan, K., Kyriazopoulou Panagiotopoulou, S., McRae, J.F., Darbandi, S.F., Knowles, D., Li, Y.I., Kosmicki, J.A., Arbelaez, J., Cui, W., Schwartz, G.B., et al. (2019) Predicting Splicing from Primary Sequence with Deep Learning. Cell, 176, 535–548.e24.

5. Sciascia, S., Roccatello, D., Salvatore, M., Carta, C., Cellai, L.L., Ferrari, G., Lumaka, A., Groft, S., Alanay, Y., Azam, M., et al. (2023) Unmet needs in countries participating in the undiagnosed diseases network international: an international survey considering national health care and economic indicators. Front Public Health, 11.

6. Graessner, H., Zurek, B., Hoischen, A. and Beltran, S. (2021) Solving the unsolved rare diseases in Europe. European Journal of Human Genetics, 29, 1319–1320.

7. Katsonis, P., Koire, A., Wilson, S.J., Hsu, T.-K., Lua, R.C., Wilkins, A.D. and Lichtarge, O. (2014) Single nucleotide variations: biological impact and theoretical interpretation. Protein Sci, 23, 1650–66.

8. Walsh, I.M., Bowman, M.A., Soto Santarriaga, I.F., Rodriguez, A. and Clark, P.L. (2020) Synonymous codon substitutions perturb cotranslational protein folding in vivo and impair cell fitness. Proc Natl Acad Sci U S A, 117, 3528–3534.

9. Dhindsa, R.S., Wang, Q., Vitsios, D., Burren, O.S., Hu, F., DiCarlo, J.E., Kruglyak, L., MacArthur, D.G., Hurles, M.E. and Petrovski, S. (2022) A minimal role for synonymous variation in human disease. Am J Hum Genet, 109, 2105–2109.

10. Shi, F., Yao, Y., Bin, Y., Zheng, C.-H. and Xia, J. (2019) Computational identification of deleterious synonymous variants in human genomes using a feature-based approach. BMC Med Genomics, 12, 12.

11. Giacoletto, C.J., Rotter, J.I., Grody, W.W. and Schiller, M.R. (2023) Synonymous Variants of Uncertain Silence. Int J Mol Sci, 24.

12. Lin, B.C., Katneni, U., Jankowska, K.I., Meyer, D. and Kimchi-Sarfaty, C. (2023) In silico methods for predicting functional synonymous variants. Genome Biol, 24, 126.

13. Jankowska, K.I., Meyer, D., Holcomb, D.D., Kames, J., Hamasaki-Katagiri, N., Katneni, U.K., Hunt, R.C., Ibla, J.C. and Kimchi-Sarfaty, C. (2022) Synonymous ADAMTS13 variants impact molecular characteristics and contribute to variability in active protein abundance. Blood Adv, 6, 5364–5378.

14. Ranganathan Ganakammal, S. and Alexov, E. (2020) An Ensemble Approach to Predict the Pathogenicity of Synonymous Variants. Genes (Basel), 11.

15. Dhindsa, R.S., Wang, Q., Vitsios, D., Burren, O.S., Hu, F., DiCarlo, J.E., Kruglyak, L., MacArthur, D.G., Hurles, M.E. and Petrovski, S. (2022) A minimal role for synonymous variation in human disease. Am J Hum Genet, 109, 2105–2109.

16. Mello, A.C., Leao, D., Dias, L., Colombelli, F., Recamonde-Mendoza, M., Turchetto-Zolet, A.C. and Matte, U. (2024) Broken silence: 22, 841 predicted deleterious synonymous variants identified in the human exome through computational analysis. Genet Mol Biol, 46, e20230125.

17. Gudkov, M., Thibaut, L. and Giannoulatou, E. (2024) Quantifying negative selection on synonymous variants. Human Genetics and Genomics Advances, 5, 100262.

18. Kovacs, E., Tompa, P., Liliom, K. and Kalmar, L. (2010) Dual coding in alternative reading frames correlates with intrinsic protein disorder. Proc Natl Acad Sci U S A, 107, 5429–34.

19. Vasu, K., Khan, D., Ramachandiran, I., Blankenberg, D. and Fox, P.L. (2022) Analysis of nested alternate open reading frames and their encoded proteins. NAR Genom Bioinform, 4.

20. Vihinen, M. (2022) When a Synonymous Variant Is Nonsynonymous. Genes (Basel), 13, 1485.

21. Ranganathan Ganakammal, S. and Alexov, E. (2020) An Ensemble Approach to Predict the Pathogenicity of Synonymous Variants. Genes (Basel), 11.

22. Zeng, Z., Aptekmann, A.A. and Bromberg, Y. (2021) Decoding the effects of synonymous variants. Nucleic Acids Res, 49, 12673–12691.

23. Buske, O.J., Manickaraj, A., Mital, S., Ray, P.N. and Brudno, M. (2015) Identification of deleterious synonymous variants in human genomes. Bioinformatics, 31, 799.

24. Zeng, Z. and Bromberg, Y. (2019) Predicting Functional Effects of Synonymous Variants: A Systematic Review and Perspectives. Front Genet, 10, 914.

25. Zeng, Z., Aptekmann, A.A. and Bromberg, Y. (2021) Decoding the effects of synonymous variants. Nucleic Acids Res, 49, 12673–12691.

26. Shi, F., Yao, Y., Bin, Y., Zheng, C.-H. and Xia, J. (2019) Computational identification of deleterious synonymous variants in human genomes using a feature-based approach. BMC Med Genomics, 12, 12.

27. Zeng, Z. and Bromberg, Y. (2019) Predicting Functional Effects of Synonymous Variants: A Systematic Review and Perspectives. Front Genet, 10, 914.

28. Mount, S.M. (1982) A catalogue of splice junction sequences. Nucleic Acids Res, 10, 459–472.

29. Will, C.L. and Luhrmann, R. (2011) Spliceosome Structure and Function. Cold Spring Harb Perspect Biol, 3, a003707–a003707.

30. Jaganathan, K., Kyriazopoulou Panagiotopoulou, S., McRae, J.F., Darbandi, S.F., Knowles, D., Li, Y.I., Kosmicki, J.A., Arbelaez, J., Cui, W., Schwartz, G.B., et al. (2019) Predicting Splicing from Primary Sequence with Deep Learning. Cell, 176, 535–548.e24.

31. Ha, C., Kim, J.-W. and Jang, J.-H. (2021) Performance Evaluation of SpliceAI for the Prediction of Splicing of NF1 Variants. Genes (Basel), 12, 1308.

32. Yeo, G. and Burge, C.B. (2003) Maximum entropy modeling of short sequence motifs with applications to RNA splicing signals. In Proceedings of the seventh annual international conference on Research in computational molecular biology. ACM, New York, NY, USA, pp. 322–331.

33. Eng, L., Coutinho, G., Nahas, S., Yeo, G., Tanouye, R., Babaei, M., Dörk, T., Burge, C. and Gatti, R.A. (2004) Nonclassical Splicing Mutations in the Coding and Noncoding Regions of the ATM Gene: Maximum Entropy Estimates of Splice Junction Strengths. Hum Mutat, 23, 67–76.

34. Gaither, J.B.S., Lammi, G.E., Li, J.L., Gordon, D.M., Kuck, H.C., Kelly, B.J., Fitch, J.R. and White, P. (2021) Synonymous variants that disrupt messenger RNA structure are significantly constrained in the human population. Gigascience, 10.

35. Davydov, E. V, Goode, D.L., Sirota, M., Cooper, G.M., Sidow, A. and Batzoglou, S. (2010) Identifying a high fraction of the human genome to be under selective constraint using GERP++. PLoS Comput Biol, 6, e1001025.

36. Cooper, G.M., Stone, E.A., Asimenos, G., NISC Comparative Sequencing Program, Green, E.D., Batzoglou, S. and Sidow, A. (2005) Distribution and intensity of constraint in mammalian genomic sequence. Genome Res, 15, 901–13.

37. Huber, C.D., Kim, B.Y. and Lohmueller, K.E. (2020) Population genetic models of GERP scores suggest pervasive turnover of constrained sites across mammalian evolution. PLoS Genet, 16, e1008827.

38. Henn, B.M., Botigué, L.R., Peischl, S., Dupanloup, I., Lipatov, M., Maples, B.K., Martin, A.R., Musharoff, S., Cann, H., Snyder, M.P., et al. (2016) Distance from sub-Saharan Africa predicts mutational load in diverse human genomes. Proc Natl Acad Sci U S A, 113, E440–9.

39. Marsden, C.D., Ortega-Del Vecchyo, D., O’Brien, D.P., Taylor, J.F., Ramirez, O., Vilà, C., Marques-Bonet, T., Schnabel, R.D., Wayne, R.K. and Lohmueller, K.E. (2016) Bottlenecks and selective sweeps during domestication have increased deleterious genetic variation in dogs. Proc Natl Acad Sci U S A, 113, 152–7.

40. Paulet, D., David, A. and Rivals, E. (2017) Ribo-seq enlightens codon usage bias. DNA Res, 24, 303–210.

41. Xu, X., Liu, Q., Fan, L., Cui, X. and Zhou, X. (2008) Analysis of synonymous codon usage and evolution of begomoviruses. J Zhejiang Univ Sci B, 9, 667–674.

42. Cutter, A.D., Wasmuth, J.D. and Blaxter, M.L. (2006) The Evolution of Biased Codon and Amino Acid Usage in Nematode Genomes. Mol Biol Evol, 23, 2303–2315.

43. Liao, X., Zhu, W., Zhou, J., Li, H., Xu, X., Zhang, B. and Gao, X. (2023) Repetitive DNA sequence detection and its role in the human genome. Commun Biol, 6, 954.

44. Zhang, Z. and Gerstein, M. (2003) Patterns of nucleotide substitution, insertion and deletion in the human genome inferred from pseudogenes. Nucleic Acids Res, 31, 5338–48.

45. Buske, O.J., Manickaraj, A., Mital, S., Ray, P.N. and Brudno, M. (2015) Identification of deleterious synonymous variants in human genomes. Bioinformatics, 31, 799.

46. Yu, G., Wang, L.-G., Han, Y. and He, Q.-Y. (2012) clusterProfiler: an R Package for Comparing Biological Themes Among Gene Clusters. OMICS, 16, 284–287.

47. Wu, T., Hu, E., Xu, S., Chen, M., Guo, P., Dai, Z., Feng, T., Zhou, L., Tang, W., Zhan, L., et al. (2021) clusterProfiler 4.0: A universal enrichment tool for interpreting omics data. The Innovation, 2, 100141.

48. Draghici, S., Khatri, P., Tarca, A.L., Amin, K., Done, A., Voichita, C., Georgescu, C. and Romero, R. (2007) A systems biology approach for pathway level analysis. Genome Res, 17, 1537–1545.

49. Wen, P., Xiao, P. and Xia, J. (2016) dbDSM: a manually curated database for deleterious synonymous mutations. Bioinformatics, 32, 1914–6.

50. Landrum, M.J., Lee, J.M., Benson, M., Brown, G., Chao, C., Chitipiralla, S., Gu, B., Hart, J., Hoffman, D., Hoover, J., et al. (2016) ClinVar: public archive of interpretations of clinically relevant variants. Nucleic Acids Res, 44, D862–8.

51. Chawla, N. V., Bowyer, K.W., Hall, L.O. and Kegelmeyer, W.P. (2002) SMOTE: Synthetic Minority Over-sampling Technique. Journal of Artificial Intelligence Research, 16, 321–357.

52. Shankar Rao Pandala (2020) Lazy Predict. https://pypi.org/project/lazypredict.

53. Chicco, D. and Jurman, G. (2020) The advantages of the Matthews correlation coefficient (MCC) over F1 score and accuracy in binary classification evaluation. BMC Genomics, 21, 6.

54. Sokolova, M., Japkowicz, N. and Szpakowicz, S. (2006) Beyond accuracy, F-score and ROC: A family of discriminant measures for performance evaluation. In AAAI Workshop - Technical Report.Vol. WS-06-06, pp. 24–29.

55. Sangkuhl, K., Römpler, H., Busch, W., Karges, B. and Schöneberg, T. (2005) Nephrogenic diabetes insipidus caused by mutation of Tyr205: a key residue of V2 vasopressin receptor function. Hum Mutat, 25, 505.

56. Schoneberg, T., Yun, J., Wenkert, D. and Wess, J. (1996) Functional rescue of mutant V2 vasopressin receptors causing nephrogenic diabetes insipidus by a co-expressed receptor polypeptide. EMBO J, 15, 1283–91.

57. Bonner, T.I., Young, A.C., Brann, M.R. and Buckley, N.J. (1988) Cloning and expression of the human and rat m5 muscarinic acetylcholine receptor genes. Neuron, 1, 403–10.

58. Römpler, H., Yu, H.-T., Arnold, A., Orth, A. and Schöneberg, T. (2006) Functional consequences of naturally occurring DRY motif variants in the mammalian chemoattractant receptor GPR33. Genomics, 87, 724–32.

59. Liebing, A.-D., Krumbholz, P. and Stäubert, C. (2023) Protocol to characterize Gi/o and Gs protein-coupled receptors in transiently transfected cells using ELISA and cAMP measurements. STAR Protoc, 4, 102120.

60. Kinsella, R.J., Kähäri, A., Haider, S., Zamora, J., Proctor, G., Spudich, G., Almeida-King, J., Staines, D., Derwent, P., Kerhornou, A., et al. (2011) Ensembl BioMarts: a hub for data retrieval across taxonomic space. Database (Oxford), 2011, bar030.

61. Karczewski, K.J., Francioli, L.C., Tiao, G., Cummings, B.B., Alföldi, J., Wang, Q., Collins, R.L., Laricchia, K.M., Ganna, A., Birnbaum, D.P., et al. (2020) The mutational constraint spectrum quantified from variation in 141, 456 humans. Nature, 581, 434–443.

62. Traag, V.A., Waltman, L. and van Eck, N.J. (2019) From Louvain to Leiden: guaranteeing well-connected communities. Sci Rep, 9.

63. Shankar Rao Pandala (2022) Lazy Predict.

64. Geurts, P., Ernst, D. and Wehenkel, L. (2006) Extremely randomized trees. Mach Learn, 63, 3–42.

65. Rentzsch, P., Witten, D., Cooper, G.M., Shendure, J. and Kircher, M. (2019) CADD: predicting the deleteriousness of variants throughout the human genome. Nucleic Acids Res, 47, D886–D894.

66. Schaafsma, G.C.P. and Vihinen, M. (2015) VariSNP, a benchmark database for variations from dbSNP. Hum Mutat, 36, 161–6.

67. Deng, Y., Gao, L., Wang, B. and Guo, X. (2015) HPOSim: an R package for phenotypic similarity measure and enrichment analysis based on the human phenotype ontology. PLoS One, 10, e0115692.

68. Gaither, J.B.S., Lammi, G.E., Li, J.L., Gordon, D.M., Kuck, H.C., Kelly, B.J., Fitch, J.R. and White, P. (2021) Synonymous variants that disrupt messenger RNA structure are significantly constrained in the human population. Gigascience, 10.

69. de Coulgeans, C.D., Silvy, M., Halverson, G., Chiaroni, J., Bailly, P. and Chapel-Fernandes, S. (2014) Synonymous nucleotide polymorphisms influence <SCP>D</SCP> ombrock blood group protein expression in <SCP>K</SCP> 562 cells. Br J Haematol, 164, 131–141.

70. Simhadri, V.L., Hamasaki-Katagiri, N., Lin, B.C., Hunt, R., Jha, S., Tseng, S.C., Wu, A., Bentley, A.A., Zichel, R., Lu, Q., et al. (2017) Single synonymous mutation in factor IX alters protein properties and underlies haemophilia B. J Med Genet, 54, 338–345.

71. Schernthaner-Reiter, M.H., Adams, D., Trivellin, G., Ramnitz, M.S., Raygada, M., Golas, G., Faucz, F.R., Nilsson, O., Nella, A.A., Dileepan, K., et al. (2016) A novel AVPR2 splice site mutation leads to partial X-linked nephrogenic diabetes insipidus in two brothers. Eur J Pediatr, 175, 727–733.

72. Pruner, I., Farm, M., Tomic, B., Gvozdenov, M., Kovac, M., Miljic, P., Soutari, N.M.H., Antovic, A., Radojkovic, D., Antovic, J., et al. (2020) The Silence Speaks, but We Do Not Listen: Synonymous c.1824C&amp;gt;T Gene Variant in the Last Exon of the Prothrombin Gene as a New Prothrombotic Risk Factor. Clin Chem, 66, 379–389.

73. Durkie, M., Cassidy, E.-J., Berry, I., Owens, M., Turnbull, C., Scott, R.H., Taylor, R.W., Deans, Z.C., Ellard, S., Baple, E.L., et al. (2024) ACGS Best Practice Guidelines for Variant Classification in Rare Disease 2024.

